# Post-acute dyslipidemia and abnormal BMI in children and adolescents with COVID-19: An EHR Cohort Study from the RECOVER Initiative

**DOI:** 10.1101/2025.09.17.25336020

**Authors:** Yuqing Lei, Ting Zhou, Bingyu Zhang, Dazheng Zhang, Huilin Tang, Jiajie Chen, Qiong Wu, Lu Li, L. Charles Bailey, Michael J. Becich, Saul Blecker, Dimitri A Christakis, Daniel Fort, Sharon J. Herring, Wenke Hwang, Amrik Singh Khalsa, Susan Kim, David M. Liebovtiz, Abu Saleh Mohammad Mosa, Suchitra Rao, Soumitra Sengupta, Xing Song, Yacob G. Tedla, Ravi Jhaveri, Caren Mangarelli, Christopher B Forrest, Yong Chen, the RECOVER Consortium

**Affiliations:** The Center for Health AI and Synthesis of Evidence (CHASE), University of Pennsylvania, Philadelphia, PA, USA; Department of Biostatistics, Epidemiology, and Informatics, University of Pennsylvania Perelman School of Medicine, Philadelphia, PA, USA; The Graduate Group in Applied Mathematics and Computational Science, School of Arts and Sciences, University of Pennsylvania, Philadelphia, PA, USA; Department of Biostatistics and Health Data Science, University of Pittsburgh, Pittsburgh, PA, USA; Applied Clinical Research Center, Children’s Hospital of Philadelphia, Philadelphia PA, USA; Department of Biomedical Informatics, University of Pittsburgh School of Medicine, Pittsburg, PA, USA; Department of Population Health, NYU Grossman School of Medicine, New York, NY, USA; Center for Child Health, Behavior and Development, Seattle Children’s Research Institute, Seattle, WA, USA; Center for Outcomes Research, Ochsner Health, New Orleans, LA, USA; Program for Maternal Health Equity, Center for Urban Bioethics, Department of Population Health and Urban Bioethics, Center for Obesity Research and Education, College of Public Health, Lewis Katz School of Medicine at Temple University, Philadelphia, PA, USA; Department of Public Health Sciences, Penn State University College of Medicine, Hershey, PA, USA; Division of Primary Care Pediatrics, Center for Child Health Equity and Outcomes Research, Abigail Wexner Research Institute, Nationwide Children’s Hospital, Columbus, OH, USA; Department of Pediatrics, College of Medicine, The Ohio State University, Columbus, OH, USA; University of California, San Francisco, Division of Rheumatology, Benioff Children’s Hospital, San Francisco, CA, USA; Department of Medicine, Northwestern University Feinberg School of Medicine, Chicago, IL, USA; Department of Biomedical Informatics and Data Science, University of Alabama at Birmingham, Heersink School of Medicine, Birmingham, AL, USA; Department of Pediatrics, University of Colorado School of Medicine and Children’s Hospital Colorado, Aurora, CO, USA; Department of Biomedical Informatics, Columbia University, New York, NY, USA; Health Management and Informatics, University of Missouri School of Medicine, Columbia, MO, USA; Division of Epidemiology, Department of Medicine, Vanderbilt University Medical Center, Nashville, TN, USA; Division of Pediatric Infectious Diseases, Ann & Robert H. Lurie Children’s Hospital of Chicago, Chicago, IL, USA; Division of Advanced General Pediatrics and Primary Care, Northwestern University Feinberg School of Medicine, Chicago, IL, USA; Department of Pediatrics, Children’s Hospital of Philadelphia, Perelman School of Medicine, University of Pennsylvania, Philadelphia, USA; Penn Medicine Center for Evidence-based Practice (CEP), Philadelphia, PA, USA; Penn Institute for Biomedical Informatics (IBI), Philadelphia, PA, USA

## Abstract

**Background:** Adults with SARS-CoV-2 infection have shown higher risks of dyslipidaemia and abnormal body mass index (BMI). Whether similar associations exist in children and adolescents is unclear.

**Method:** We did a retrospective cohort study using the RECOVER paediatric Electronic Health Record (EHR) datasets from 25 US children’s hospitals, covering March 2020 to September 2023. For dyslipidaemia analyses, we included 384,289 patients aged 0–21 years with at least 6 months of follow-up and 1,080,413 COVID-19-negative controls. For BMI analyses, we included 285,559 patients aged 2–21 years and 817,315 controls. Documented infection was defined as a positive PCR, serology, or antigen test, or a clinical diagnosis of COVID-19 or post-acute sequelae of SARS-CoV-2. Outcomes were new diagnoses of dyslipidaemia, defined by laboratory thresholds for total cholesterol, triglycerides, LDL cholesterol, HDL cholesterol, and non-HDL cholesterol, and abnormal BMI (BMI-for-age ≥95th percentile at ages 2–19 years or BMI ≥30 kg/m² at ages 19–21 years). Adjusted relative risks (aRRs) were estimated using propensity score-stratified Poisson regression. Sensitivity analyses included empirical calibration with negative control outcomes and stratification by baseline obesity.

**Interpretation:** Children and adolescents with documented COVID-19 were associated with higher risks of new-onset dyslipidaemia and abnormal BMI in the post-acute period compared with COVID-19-negative peers. Associations were consistent across lipid fractions, remained after empirical calibration, and were similar after accounting for baseline obesity.

**Research in context:** 

**Evidence before this study:** Adults with SARS-CoV-2 infection have been reported to develop dyslipidaemia and abnormal body mass index (BMI) after the acute phase, raising concerns about long-term metabolic health. In children and adolescents, evidence has been scarce. Available studies are small, cross-sectional, or based mainly on diagnosis codes, with few incorporating laboratory lipid values or age-specific BMI thresholds against contemporaneous controls. The risk of post-acute dyslipidaemia and BMI abnormalities in paediatric populations therefore remains uncertain.

**Added value of this study:** Using the Researching COVID to Enhance Recovery (RECOVER) electronic health record (EHR) database from 25 US hospitals, we examined more than 1.6 million children and adolescents with at least 6 months of follow-up. Outcomes were defined using laboratory lipid panels and age-specific BMI measures. With propensity score stratification across hundreds of covariates and calibration using negative control outcomes, documented COVID-19 was associated with higher adjusted risks of abnormal HDL cholesterol, LDL cholesterol, total cholesterol, triglycerides, and BMI. Associations were consistent across sensitivity analyses and stratified by baseline obesity.

**Implications of all the available evidence:** Together with findings from adult studies, our results indicate that metabolic sequelae after SARS-CoV-2 infection are also relevant in paediatric populations. Children and adolescents with documented COVID-19 were more likely to develop dyslipidaemia and abnormal BMI in the early post-acute phase. These findings support routine lipid and BMI monitoring in paediatric follow-up care, which could enable earlier identification of metabolic dysfunction and guide preventive strategies for long-term cardiometabolic health.

## Introduction

Children and adolescents infected with SARS-CoV-2 generally experience milder acute illness than adults, yet accumulating evidence suggests substantial post-acute health consequences^1^. Furthermore, risks for post-acute sequelae vary by prior infection and reinfection, vaccine status, sociodemographic factors, and socioeconomic status^2–5^. Recent cohort studies have reported increased risks of cardiovascular, gastrointestinal, renal, and neuropsychiatric sequelae following pediatric COVID-19^6–9^, suggesting that post-acute sequelae of SARS-CoV-2 (PASC) extend beyond respiratory symptoms in young populations.

The metabolic health of children and adolescents represents a critical public health concern, particularly as childhood obesity and dyslipidemia are established risk factors for various health issues, such as coronary artery disease, metabolic dysfunction-associated steatosis liver disease, and type 2 diabetes^10,11^. Pre-pandemic data from the National Health and Nutrition Examination Survey (NHANES) indicated that 21% of U.S. children and adolescents aged 6-19 had at least one abnormal cholesterol measure, 32% had a high body mass index (BMI)^12^. While lifestyle disruptions during the pandemic (e.g., decreased physical activity, dietary changes) have contributed to rising rates of pediatric abnormal BMI and dyslipidemia, the extent to which these trends reflect direct post-infectious effects of COVID-19 remains uncertain. Recent studies found associations between pre-infection BMI and post-acute COVID-19 outcomes^13,14^, yet whether COVID-19 infection is associated with subsequent abnormal BMI and dyslipidemia in children has not been comprehensively evaluated.

In adults, one study leveraging national health databases from the US Department of Veterans Affairs had found an increased risk of incident dyslipidemia in COVID-19 survivors, with effects persisting well beyond one year^15^. Although emerging evidence suggests that children may also experience metabolic disturbances related to inflammatory responses to SARS-CoV-2^16^, the pediatric data remain scarce: existing studies are often constrained by short follow-up or incomplete lipid measurements^17,18^, leaving the metabolic consequences of COVID-19 in children and adolescents largely undefined. This knowledge gap is particularly concerning given that metabolic abnormalities acquired during childhood often persist into adulthood, potentially amplifying lifetime cardiovascular risk^19,20^.

To fill this gap, we conducted a retrospective cohort study using the Researching COVID to Enhance Recovery (RECOVER) electronic health records (EHR) database, involving 25 US children’s hospitals and health institutions, to explore whether documented SARS-CoV-2 infection was associated with increased risks of dyslipidemia and abnormal BMI among children and adolescents, during the post-acute period (28-179 days). By analyzing 384,289 COVID-19-positive patients for dyslipidemia outcomes and 285,559 for BMI outcomes, with corresponding COVID-19-negative controls, this investigation represents the largest comprehensive assessment of post-COVID-19 metabolic sequelae in the pediatric population. Our findings aim to inform clinical monitoring strategies and preventive interventions for the millions of children recovering from COVID-19 worldwide.

## Methods

### Study Design and Data Sources

This retrospective cohort study constitutes human subject research. Institute Review Board (IRB) approval was obtained under Biomedical Research Alliance of New York (BRANY) protocol #21-08-508. As part of the BRANY IRB process, the protocol has been reviewed in accordance with the Strengthening the Reporting of Observational Studies in Epidemiology (STROBE) reporting guideline^21^. The BRANY waived the need for consent and HIPAA authorization. This study is affiliated with the NIH RECOVER Initiative (https://recovercovid.org/), a project dedicated to understanding the enduring impacts of COVID-19. The dataset comprises information from 25 contributing sites from March 2020 through September 2023. Institutional review boards at participating sites approved the protocol with waiver of informed consent because only de-identified data were used.

### Participants

There are two parallel cohorts with at least 6 months of follow-up: (1) children and adolescents aged 0-21 years^22^ for dyslipidemia analysis, and (2) those aged 2-21 years for abnormal BMI analysis. For the COVID-19-positive group, the index date was defined as the earliest documented evidence of SARS-CoV-2 infection—a positive polymerase-chain-reaction (PCR), serology, antigen tests, or a documented diagnosis of COVID-19 or PASC–thereby anchoring follow-up at the actual onset of acute illness. We contrasted these patients against a COVID-19-negative control group without any record of SARS-CoV-2 infection who had at least one negative COVID-19 test during the same calendar period. For the control group, the index date was randomly assigned based on the empirical distribution of index dates in COVID-19 patients, to ensure comparable temporal risk windows. We excluded patient who did not have any clinical visits in the 24 months (729 days) to 7 days before their index date (baseline period) or between 28 and 179 days after their index date (follow-up period). Additionally, patients with a history of dyslipidemia or abnormal BMI during the baseline period were further excluded from the corresponding analyses. For abnormal BMI study cohorts, we further excluded patients who took weight-modifying medications as well as those with associated abnormal conditions at the baseline period (refer to Supplementary **Section S1C, S2** for further details).

### Defining Dyslipidemia Outcomes

We adapted our lipid thresholds from Xu et al.’s work^15^ and established pediatric guidelines^20,23^. Dyslipidemia outcomes consisted of the following abnormal lipid laboratory results: abnormal total cholesterol (TC): ≥ 200 mg/dL; abnormal triglycerides (TG) ≥ 100 mg/dL (ages 0-9 years); ≥ 130 mg/dL for (ages 10-19 years), ≥ 150 mg/dL for (ages 20-21 years); abnormal low-density lipoprotein (LDL) cholesterol: ≥ 130 mg/dL; abnormal high-density lipoprotein (HDL) cholesterol: < 40 mg/dL; abnormal non-HDL cholesterol: ≥ 145 mg/dL. We assessed incidents of each dyslipidemia outcome during the post-acute phase of COVID-19 (28-179 days after index) among those without any history of dyslipidemia in the two years preceding the index. The composite of any dyslipidemia outcome—“any abnormal lipid laboratory result”—was defined as the first occurrence of any of the above thresholds during the study period.

### Defining Abnormal BMI Outcome

Abnormal BMI outcomes were determined based on age-appropriate BMI thresholds from recent pediatric growth and treatment guidelines^24–27^. For ages 2-18 years: BMI-for-age ≥ 95th percentile (based on CDC groth charts, operationalized using BMI z-scores), For ages 19-21 years: BMI ≥ 30 kg/m^2^. The incidence of abnormal BMI outcome was ascertained over the same follow-up window (28-179 days after index), excluding individuals with any history of BMI above the threshold during the two-year baseline period.

### Covariates

We adjusted for a comprehensive set of baseline characteristics to control for confounding and ensure balanced comparison between SARS-CoV-2-exposed and unexposed children. The demographic characteristics included: patient age at cohort entry (years), sex (female/male), race and ethnicity (Non-Hispanic White [NHW], Non-Hispanic Black [NHB], Hispanic, Asian American/Pacific Islander [AAPI], Multiple, Other/Unknown). The temporal and site factors included the year-month of cohort entry (from March 2020 to September 2023) and the study site. The healthcare utilization measures included: the number of inpatient, outpatient, and emergency department visits; the number of unique medications or prescriptions (0, 1, 2, ≥3); the number of COVID-19 negative tests 24 months to 7 days before the cohort entry date (0, 1, 2, ≥3). Chronic comorbidities were assessed using the Pediatric Medical Complexity Algorithm (PMCA, no chronic condition, non-complex chronic condition, complex chronic condition)^28,29^, and a list of pre-existing chronic conditions^30,31^ (see Supplementary **Section S1.D** for more details).

For the dyslipidemia analysis, we further controlled for obesity status during the baseline period and for whether each patient completed corresponding lipid lab tests at the baseline period^32,33^ (see Supplementary **Section S1.D** for more details).

### Statistical Analysis

To ensure only incident cases were captured, we first implemented a 24-month baseline washout period by excluding any patients with pre-existing abnormalities of lipid or BMI. We then compared the incidences of new-onset dyslipidemia or abnormal BMI outcomes between COVID-19-positive and negative control cohorts, by calculating the number of patients developing the specified outcome during the post-acute window (28-179 days after index) divided by the total number of patients in each cohort. Outcomes with an overall incidence below 0.1% were excluded a priori to prevent model instability^34^. Distributions of preference scores^35^— a transformation of propensity scores that accounts for prevalence differences between groups— were examined to assess empirical equipoise (Supplementary **Section S2.C**). To mitigate potential confounding due to imbalanced baseline between comparable groups, we estimated each patient’s propensity score, the probability of being in the COVID-19-positive group given covariates, using a multivariate logistic regression model that included pre-specified covariates as the independent variables. Patients were then stratified into six propensity-score strata, and within each stratum, we modeled each outcome via Poisson regression, incorporating the stratum identifier as a covariate, to estimate adjusted risk ratios (aRRs) to quantify the overall risk^36^. The balance was assessed via standardized mean difference (SMD) for each covariate. A difference of ≤ 0.1 was considered indicative of acceptable balance^37^.

### Sensitivity Analysis

Recognizing that unmeasured factors may still bias our aRR estimates, we performed two complementary sensitivity checks. First, we conducted empirical calibration^38–40^ using 36 negative control outcomes (NCOs)—conditions known a priori not to be causally associated with COVID-19—thereby quantifying and correcting any residual systematic error in the risk estimate. Second, for the dyslipidemia analysis, we stratified by baseline obesity status to assess potential differential effects.

## Results

### Cohort Identification

We included 384,289 COVID-19-positive children and adolescents for the dyslipidemia analysis (mean [SD] entry age, 8.6 [6.5] years) and 285,559 for the abnormal BMI analysis (10.5 [5.5] years). The COVID-19-negative control cohorts comprised 1,080,413 patients (7.9 [6.2] years) for dyslipidemia and 817,315 patients (9.5 [5.5] years) for abnormal BMI (**Table 1a,b**). **Figure 1** illustrates an overview of the participant selection process for both COVID-19-positive and COVID-19-negative patients with eligible criteria in the dataset.

**Figure 1.**
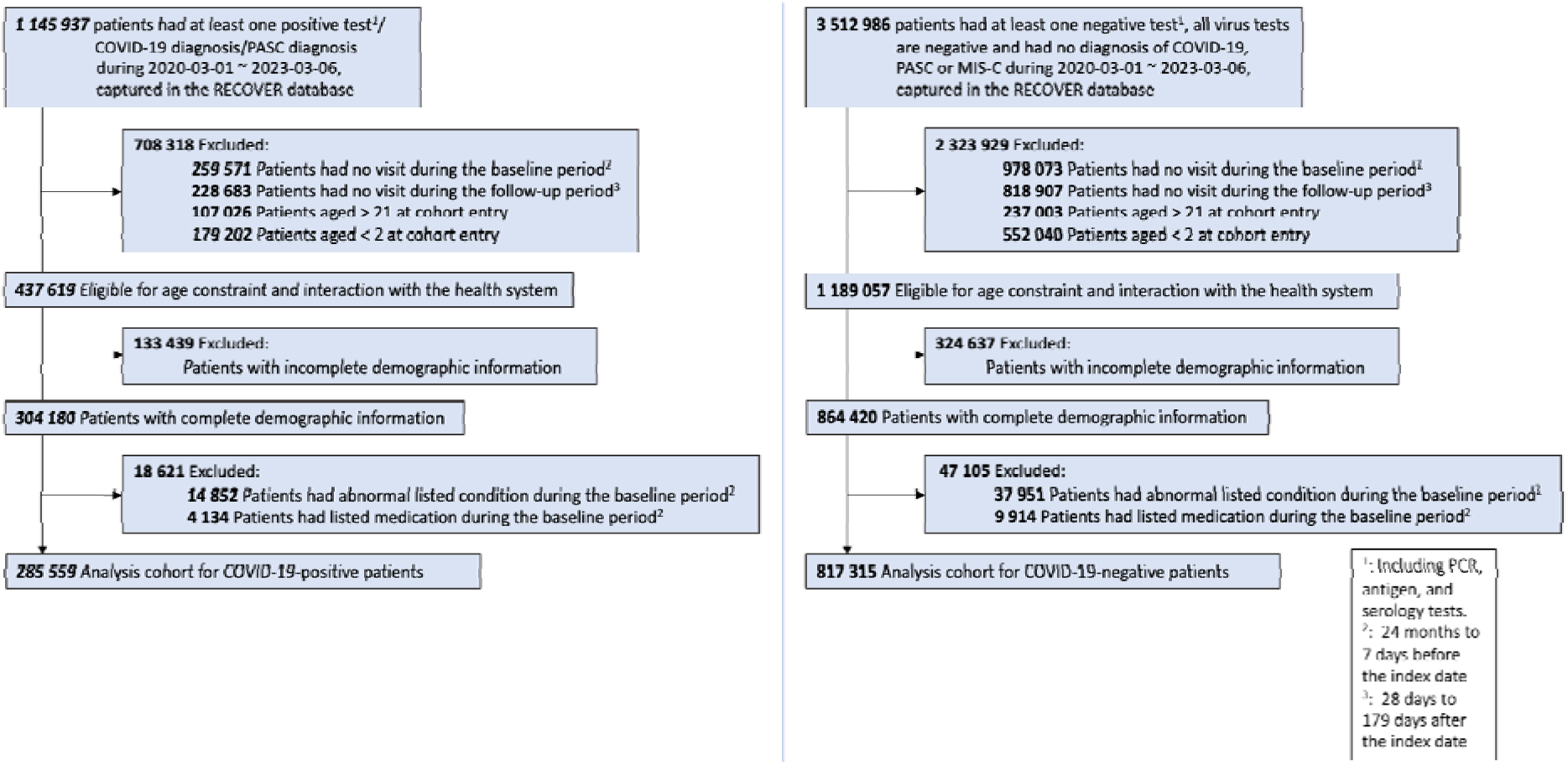
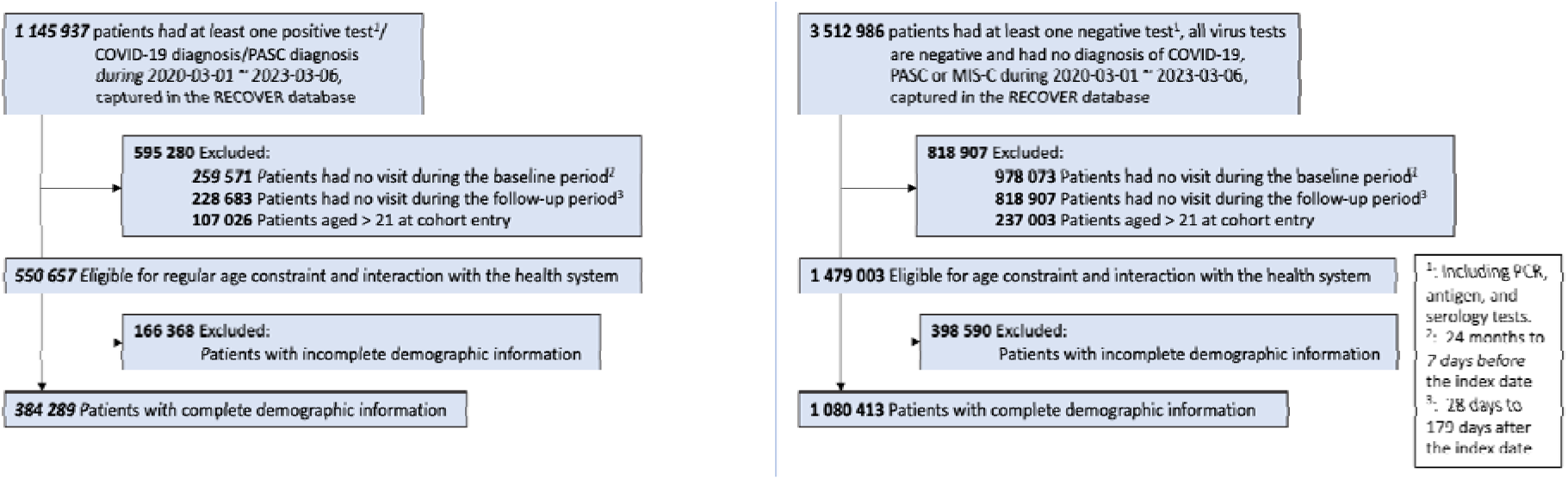
a. Selection of participants for both COVID-19-positive and COVID-19-negative patients for abnormal BMI outcome b. Selection of participants for both COVID-19-positive and COVID-19-negative

**Table 1.**
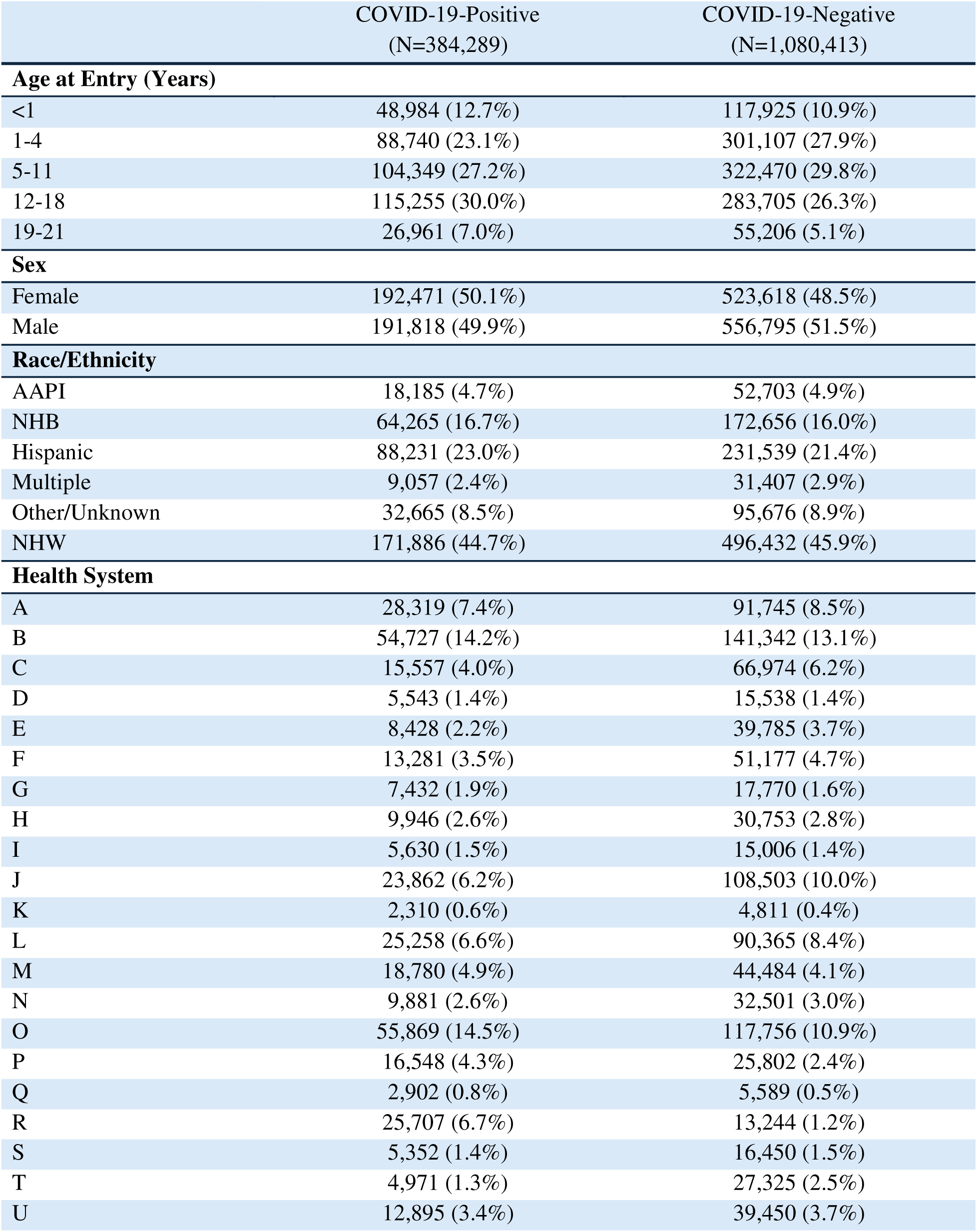
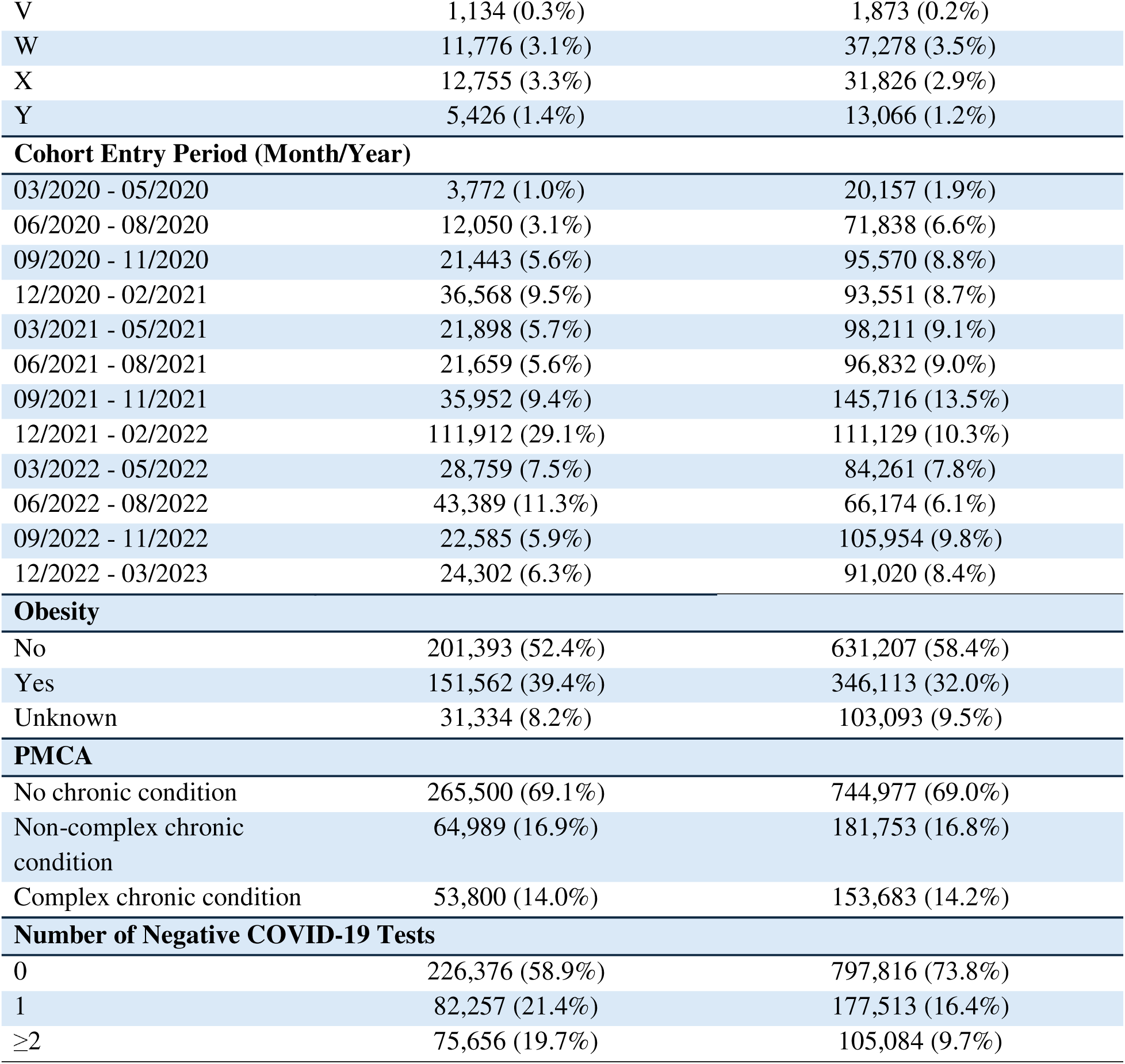
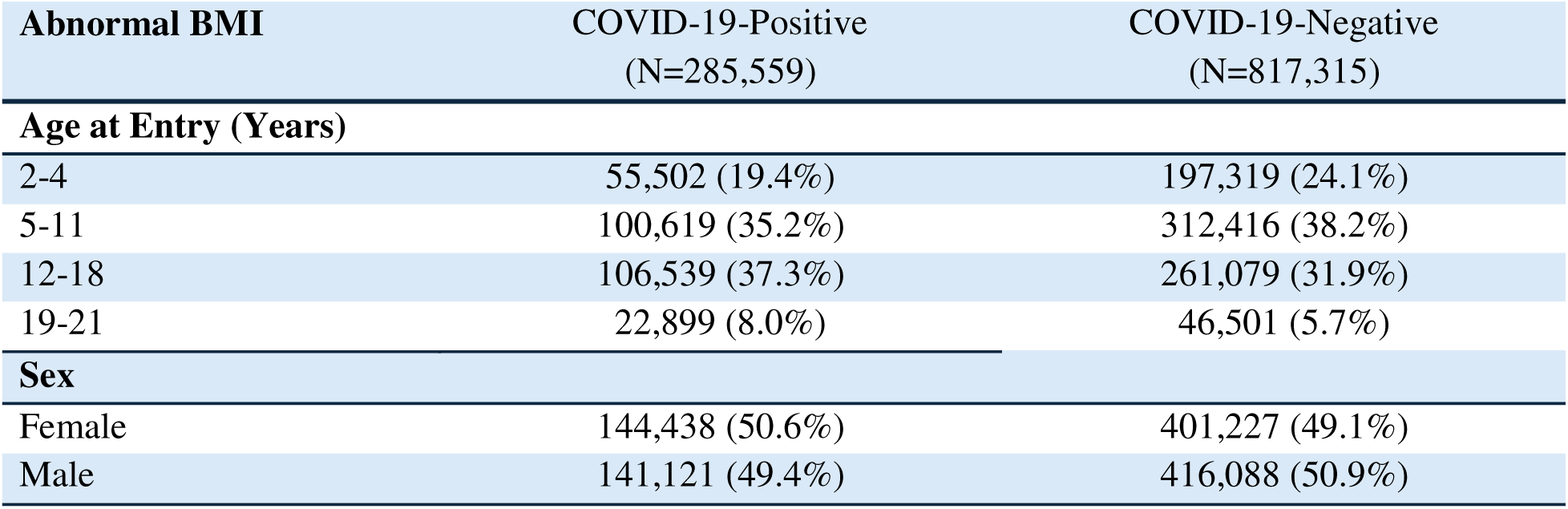
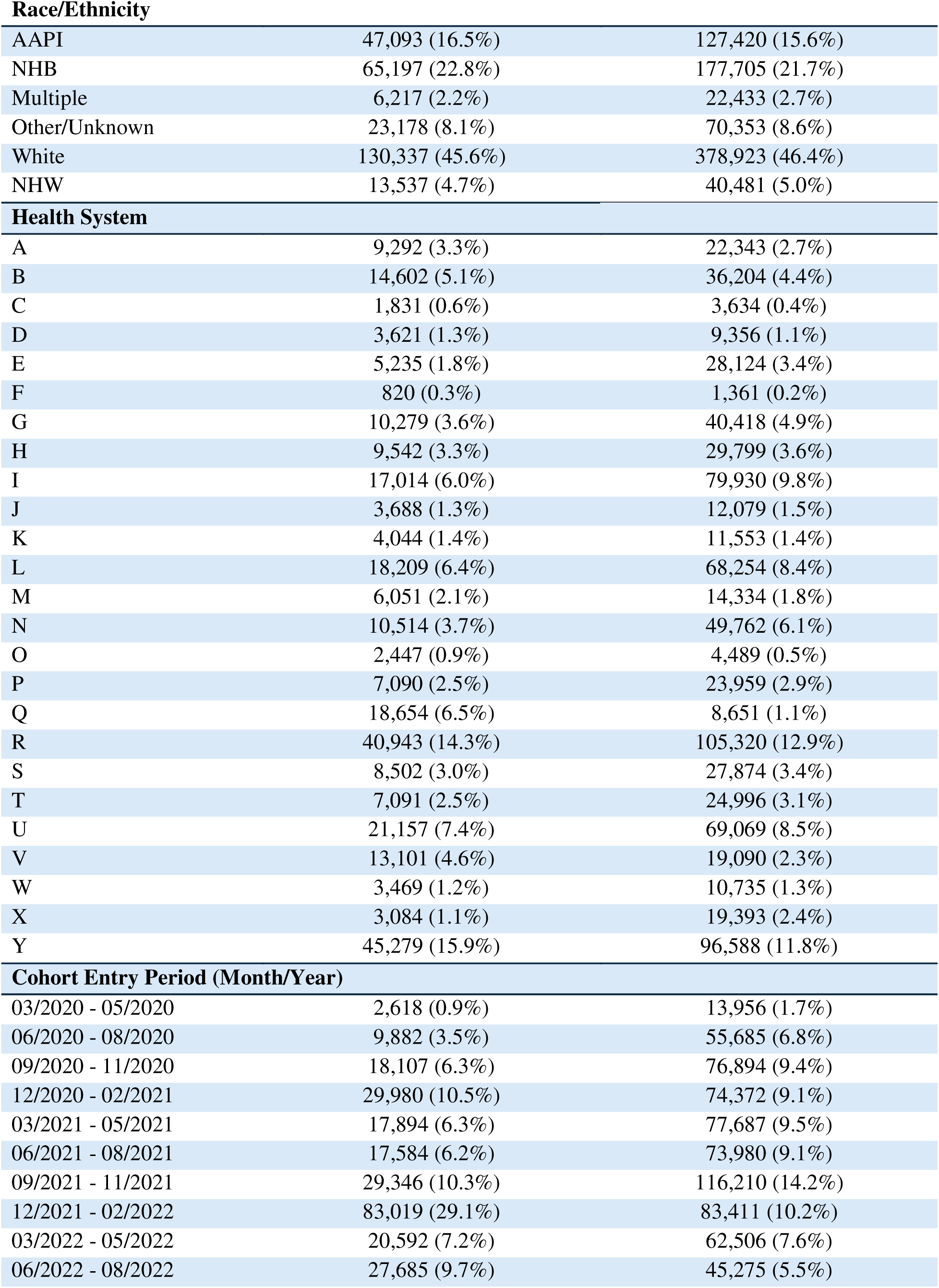
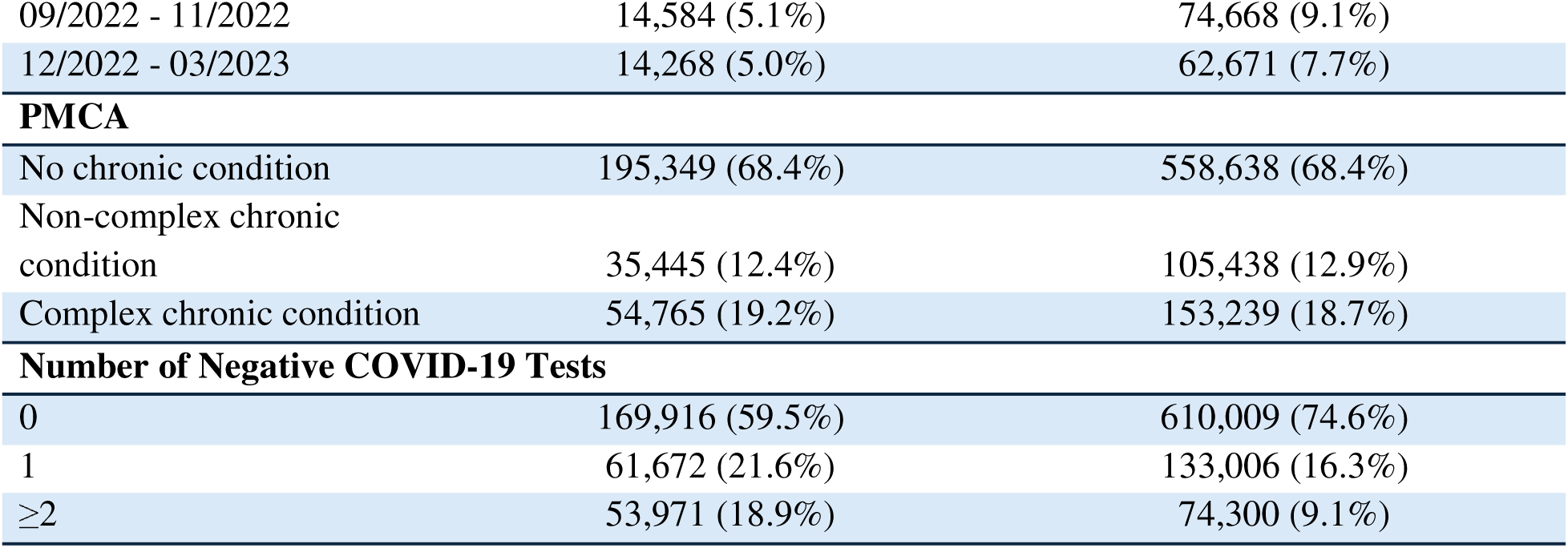
a. Baseline characteristics of patients for the dyslipidemia analysis. b. Baseline characteristics of patients for the abnormal BMI analysis.

### Incidence of Post-Acute Abnormal Outcomes for both COVID-19 and Control Groups

The incidence of post-acute composite dyslipidemia (“any abnormal lipid result”) was 0.9% in the COVID-19-positive group compared with 0.7% in COVID-19-negative controls (**Table 2**). Across individual lipid measures, incidence in the COVID-19-positive group ranged from 0.2% for LDL abnormalities to 0.6% for low HDL, compared with 0.1% and 0.4% in the COVID-19-negative group, respectively. For abnormal BMI, post-acute incidence was 6.0% among those with prior COVID-19 versus 4.7% in controls.

**Table 2.** Raw incidence (in %), calculated as the absolute number of patients with outcome during the post-acute phase divided by the number of total patients that did not have the specific outcome during the baseline period.

| Dyslipidemia Outcomes | COVID-19-Positive<br>(N=384,289) | COVID-19-Negative<br>(N=1,080,413) |
| --- | --- | --- |
| **Incident abnormal lipid laboratory results** |  |  |
| Abnormal HDL Cholesterol | 0.576 (2,164/375,603) | 0.414 (4,395/1,061,721) |
| Abnormal LDL Cholesterol | 0.197 (751/381,401) | 0.149 (1,598/1,074,385) |
| Abnormal Non-HDL Cholesterol | 0.111 (425/382,449) | 0.088 (946/1,077,298) |
| Abnormal TC | 0.268 (1,020/380,069) | 0.211 (2,260/1,071,771) |
| Abnormal TG | 0.491 (1,851/376,655) | 0.342 (3,639/1,064,911) |
| **Composite dyslipidemia outcomes** |  |  |
| Any abnormal lipid lab results | 0.945 (3,490/369,233) | 0.691 (7,254/1,049,067) |
| **Abnormal BMI Outcomes** | **COVID-19-Positive**<br>**(N=285,559)** | **COVID-19-Negative**<br>**(N=817,315)** |
| Abnormal BMI | 5.954 (10,689/179,518) | 4.743 (27,967/589,622) |

†Abnormal HDL Cholesterol: high-density lipoprotein (HDL) cholesterol: < 40 mg/dL; Abnormal LDL Cholesterol: low-density lipoprotein (LDL) cholesterol: ≥ 130 mg/dL; Abnormal Non-HDL Cholesterol: non-HDL cholesterol: ≥ 145 mg/dL; Abnormal TC: total cholesterol (TC): ≥ 200 mg/dL; Abnormal TG: triglycerides (TG) ≥ 100 mg/dL for ages 0-9 years, ≥ 130 mg/dL for ages 10-19 years, ≥ 150 mg/dL for ages 20-21 years. Any abnormal lipid lab results: the first occurrence of any of the above thresholds during the study period; Abnormal BMI: BMI z-score ≥ 95th percentile for ages 2-18 years, BMI ≥ 30 kg/m<sup>2</sup> for ages 19-21 years.

### Adjusted Relative Risk (aRR) of Post-Acute Abnormal Outcomes

After propensity score stratification, the documented SARS-CoV-2 infection was significantly associated with higher risks of most lipid abnormalities at the post-acute phase (**Figure 2**). aRRs were 1.24 (95% CI 1.18-1.31) for abnormal HDL cholesterol levels, 1.19 (95% CI 1.08-1.30) for abnormal LDL cholesterol levels, 1.14 (95% CI 1.06-1.24) for total cholesterol levels, and 1.28 (95% CI 1.20-1.36) for abnormal triglyceride levels. Non-HDL cholesterol levels showed a non-significant trend (aRR 1.03, 95% CI 0.92-1.17). The composite of any abnormal lipid result outcome yielded an aRR of 1.23 (95% CI 1.18-1.29). For abnormal BMI outcome, the COVID-19-positive patients were associated with a 15% higher risk than COVID-19-negative controls, with an aRR of 1.15 (95% CI 1.12-1.18).

**Figure 2.**
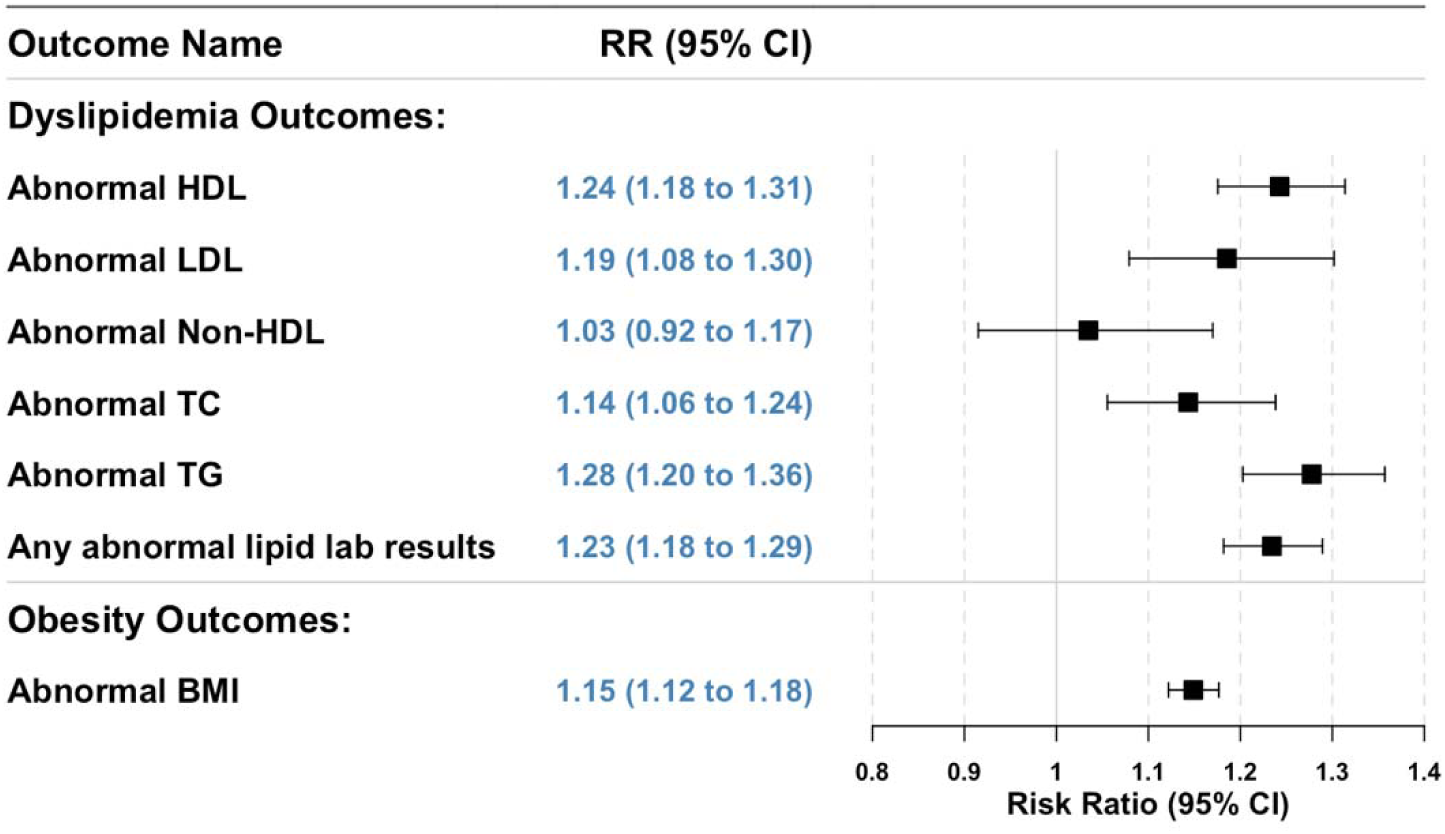
Adjusted Relative Risk (aRR) of incident post-acute COVID-19 dyslipidemia outcomes and abnormal BMI outcomes compared with the COVID-19-negative cohort. *†Abnormal HDL Cholesterol: high-density lipoprotein (HDL) cholesterol: < 40 mg/dL; Abnormal LDL Cholesterol: low-density lipoprotein (LDL) cholesterol:* ≥ *130 mg/dL; Abnormal Non-HDL Cholesterol: non-HDL cholesterol:* ≥ *145 mg/dL; Abnormal TC: total cholesterol (TC):* ≥ *200 mg/dL; Abnormal TG: triglycerides (TG)* ≥ *100 mg/dL for ages 0-9 years,* ≥ *130 mg/dL for ages 10-19 years,* ≥ *150 mg/dL for ages 20-21 years. Any abnormal lipid lab results: the first occurrence of any of the above thresholds during the study period; Abnormal BMI: BMI z-score*≥ *95t percentile for ages 2-18 years, BMI* ≥ *30 kg/m*^2^ *for ages 19-21 years*.

### Sensitivity Analysis

Empirical calibration using 36 NCOs produced point estimates very close to the primary aRRs but with modestly wider confidence intervals, suggesting minimal residual bias (Detailed results are provided in Supplementary **Section S3**). Stratification by baseline obesity status yielded consistent aRRs across subgroups, indicating that pre-existing adiposity did not materially modify our primary findings (Detailed results are provided in Supplementary **Section S4**).

## Discussion

By leveraging 25 longitudinal EHR datasets, we found that children and adolescents with documented COVID-19 infection were associated with modestly increased risks of incident dyslipidemia and abnormal BMI among children and adolescents during the 28-179-day post-acute phase. These findings represent the first large-scale evidence of post-COVID metabolic associations in the pediatric population, underscoring that children and adolescents may experience persistent metabolic consequences despite typically milder acute illness.

Our results extend observations from adult studies^15,41,42^ to the younger age group. The Veterans Affairs cohort study has shown an increased risk of incident dyslipidemia in adult COVID-19 survivors, with hazard ratios ranging from 1.2 to 1.27 for different lipid abnormalities^15^. Our pediatric findings demonstrate lower but predominantly significant effect sizes (aRR ranging from 1.04 to 1.28), suggesting that while children may have protective factors, they remain susceptible to dyslipidemia sequelae of SARS-CoV-2 infection. Similar patterns emerged for abnormal BMI associations. These BMI findings align with documented pandemic-related weight gain in children^43^, though our results suggest that COVID-19 infection itself may contribute additional metabolic risk beyond broader pandemic lifestyle effects.

Several mechanisms may explain these associations. SARS-CoV-2 may disrupt lipid homeostasis either through angiotensin-converting enzyme 2 (ACE2) receptor-mediated signaling^44–46^ or directly via its spike protein, both of which contribute to the upregulation of lipid synthesis pathways and metabolic dysregulation. Infection also triggers systemic inflammation, notably elevations in interleukin-6 and tumor necrosis factor–alpha, which have been associated with acute dyslipidemia, including reduced HDL cholesterol and elevated triglycerides^47,48^, likely through hepatic lipid metabolism disruption. These biological mechanisms, compounded by pandemic-related routine behavioral disruptions—such as school closures, reduced physical activity, increased screen time, and changes in dietary patterns—may collectively contribute to post-infection effects in dyslipidemia and abnormal BMI, particularly in vulnerable pediatric populations.

While individually modest, our findings may have substantial population-level implications given widespread pediatric COVID-19 exposure. Childhood metabolic abnormalities often persist into adulthood, potentially amplifying lifetime cardiovascular risk^10,11,19,20^. These considerations support incorporating enhanced metabolic monitoring into post-COVID pediatric follow-up care.

### Strengths and Limitations

This study has several strengths. First, it leveraged the RECOVER EHR consortium across 25 U.S. children’s hospitals to assemble the largest and most heterogeneous cohort to date for examining post-acute abnormalities of lipid and BMI in youth. Second, inclusion of contemporaneous, COVID-19 negative controls and rigorous propensity score stratification across more than hundreds of covariates minimized confounding. Third, the use of laboratory based definitions for lipid abnormalities and objectively measured BMI reduced misclassification compared with diagnosis codes alone. Fourth, multiple sensitivity analyses, including empirical calibration with 36 NCOs and stratified analyses by baseline obesity, further reinforced the reliability of our results. Finally, consistent effect estimates across individual lipid subtypes and composite outcomes underscore the clinical relevance of our findings.

However, several limitations merit consideration. First, documented SARS-CoV-2 infections may be under-ascertained if testing occurred outside the captured EHR network or among asymptomatic children, while minor differential misclassification could be corrected^49^. Second, although we balanced over hundreds of covariates, unmeasured factors—diet, physical activity, sleep patterns, and socioeconomic status—may still confound the associations; our NCO calibration provides some correction, but residual bias cannot be excluded^50^. Third, indication bias may also be present because lipid assessments are not routinely conducted in pediatric practice; it is also possible that children with pre-existing obesity or more severe symptoms were more likely to present for clinical evaluation and undergo laboratory testing, which could increase the likelihood of detecting lipid abnormalities in these groups. To address this, we further included a baseline indicator for participating in lipid tests in our covariates set. All confounders were balanced after propensity score stratification, with an SMD of less than 0.1 (Supplementary **Section S2.C**). Additionally, we excluded children receiving COVID-specific treatments but could not fully account for other medications (e.g., corticosteroids) that influence metabolic outcomes. Our analysis focused on early post-acute changes (28-179 days), leaving longer-term trajectories unclear. Finally, as an observational study, causality cannot be definitively established, although the consistency across sensitivity analyses strengthens our inference that SARS-CoV-2 infection contributes to early dyslipidemia and BMI abnormalities.

## Conclusions

Children and adolescents with documented COVID-19 infection are associated with modestly increased risks of new-onset dyslipidemia and abnormal BMI during the post-acute phase. These findings support integrating enhanced metabolic monitoring into routine post-COVID follow-up care to enable early detection and intervention. Future research should explore the underlying biological mechanisms, longer-term metabolic trajectories, and effective preventive strategies to mitigate potential cardiovascular and metabolic complications in pediatric COVID-19 survivors.

## Data Availability

Due to data use agreements and patient privacy consideration, theses data are not publicly available. Access to PEDSnet data may be granted to qualified investigators through a formal proposal process and with approval from participating institutions and the PEDSnet Data Coordinating Center.

## Acknowledgments

This study is part of the NIH Researching COVID to Enhance Recovery (RECOVER) Initiative, which seeks to understand, treat, and prevent the post-acute sequelae of SARS-CoV-2 infection (PASC). For more information on RECOVER, visit https://recovercovid.org/. We would like to thank the National Community Engagement Group (NCEG), all patients, caregivers, and community Representatives, and all the participants enrolled in the RECOVER Initiative. We would like to thank the patient representatives Christine Maughan, Etienne Carignan and Teresa Akintonwa for their helpful suggestions and comments. We also extend our sincere appreciation to Dr. Thao Ly Tam Phan Vo, MD, MPH (Associate Professor of Pediatrics and Research Scientist at Nemours Children’s Hospital, Delaware), for her expertise and invaluable clinical input in defining outcomes and shaping the study protocol.

## Source of Funding

This research was funded by the NIH Agreement OTA OT2HL161847-01 as part of the Researching COVID to Enhance Recovery (RECOVER) research Initiative. The research reported in this publication was conducted using PEDSnet, A Pediatric Clinical Research Network. PEDSnet has been developed with funding from the Patient-Centered Outcomes Research Institute (PCORI); PEDSnet’s participation in PCORnet is funded through PCORI award RI-CHOP-01-PS10. This publication includes data from the following PEDSnet institutions: Cincinnati Children’s Hospital Medical Center, Children’s Hospital of Philadelphia, Children’s Hospital Colorado, Columbia University Irving Medical Center, Ann & Robert H. Lurie Children’s Hospital of Chicago, University of Michigan, University of Missouri, Montefiore, Medical University of South Carolina (MUSC), Nationwide Children’s Hospital, University of Nebraska Medical Center, Nemours Children’s Health, Northwestern University, New York University School of Medicine, OCHIN, Inc., Ochsner Health System, Ohio State University, University of Pittsburgh/UPMC, Penn State Health and College of Medicine, Seattle Children’s Hospital, Stanford University, University of Temple, University of California San Francisco, Vanderbilt University Medical Center, Weill Cornell Medical College. Data was extracted from the RECOVER Database Version s10. PaTH is a Network Partner in PCORnet® which has been developed with funding from the Patient-Centered Outcomes Research Institute® (PCORI®) and is a data provider to RECOVER. PaTH’s participation in PCORnet is funded through PCORI Award RI-PITT-01-PS1.

## Disclosures

This content is solely the responsibility of the authors and does not necessarily represent the official views of the RECOVER Initiative, the NIH, or other funders. **The views, statements, and opinions presented in this publication are solely the responsibility of the author(s) and do not necessarily represent the views of other organizations participating in, collaborating with, or funding PCORnet® or of the Patient-Centered Outcomes Research Institute® (PCORI®).** Dr Bailey has received grants from Patient-Centered Outcomes Research Institute, Dr. Rao reports prior grant support from GSK and Biofire and is a consultant for Sequiris, Dr. Jhaveri is a consultant for AstraZeneca, Seqirus, Dynavax, receives an editorial stipend from Elsevier and Pediatric Infectious Diseases Society and royalties from Up To Date/Wolters Kluwer.

All authors have no conflicts of interest to report.

## Notes

### Competing Interest Statement

The authors have declared no competing interest.

### Author Declarations

The Biomedical Research Alliance of New York (BRANY) Institutional Review Board (protocol #21-08-508) reviewed this study and granted ethical approval with waiver of informed consent and HIPAA authorization, given the use of de-identified electronic health record data.

