## Supplementary material for "Post-acute dyslipidemia and abnormal BMI in children and adolescents with COVID-19: An EHR Cohort Study from the RECOVER Initiative": Supplymentary

### Section S1 Supplemental Methods

#### A. Description of electronic health records (EHR) data

The real-world data utilized in our analysis is derived from electronic health records (EHRs), covering a wide range of healthcare interaction information routinely collected and stored by hospitals. This includes clinical data such as diagnoses and treatments, laboratory and test results, and administrative data including patient demographics and billing information. The hospital-based EHR data from the Researching COVID to Enhance Recovery (RECOVER) Initiative COVID-19 Database served as the basis for defining and determining exposure, outcomes, and covariates. Unlike General Practitioner (GP) data, self-reported data, or external data sources, our study used the structured, standardized EHR entries made by healthcare providers within hospital settings. EHR data provides a more detailed and integrated view of a patient's health status, medical history, and healthcare interactions across various providers and settings.

#### B. RECOVER population and generalizability

The National Institutions of Health (NIH) launched the new RECOVER initiative in 2021 to leverage electronic health record (EHR) data to better identify and characterize patients with post-acute sequelae of SARS-CoV-2 infection (PASC). RECOVER obtains EHRs from three large national healthcare networks within the United States, covering regional catchment areas across 41 states. These networks collectively hold the EHRs of over 60 million patients, including records from more than 7 million individuals who have been affected by COVID-19. RECOVER collaborates with the National Institutes of Health's (NIH) All of Us Research Program, which contributes additional health records to this vast database. Together, these sources comprise one of the world's largest collections of EHRs.

In our study, participating institutions in this study included: Cincinnati Children’s Hospital Medical Center, Children’s Hospital of Philadelphia, Children’s Hospital Colorado, Columbia University Irving Medical Center, Ann & Robert H. Lurie Children's Hospital of Chicago, University of Michigan, University of Missouri, Montefiore, Medical University of South Carolina (MUSC), Nationwide Children’s Hospital, University of Nebraska Medical Center, Nemours Children’s Health, Northwestern University, New York University School of Medicine, OCHIN, Inc., Ochsner Health System, Ohio State University, University of Pittsburgh/UPMC, Penn State Health and College of Medicine, Seattle Children’s Hospital, Stanford University, University of Temple, University of California San Francisco, Vanderbilt University Medical Center, Weill Cornell Medical College. For this study, we used the s10 version of the data, collected till September 2023.

#### C. Cohort definition and observation windows

We assembled two parallel pediatric cohorts (dyslipidemia and abnormal BMI) from March 2020 through September 2023. Entry criteria and observation windows were defined as follows:

- **Age at Index**
  - Dyslipidemia cohorts: 0–21 years
  - Abnormal-BMI cohorts: 2–21 years
- **Index Date**
  - **COVID-19 Positive Cohort**: Earliest evidence of SARS-CoV-2 infection: positive PCR, antigen, or serology test; or documented diagnosis of COVID-19 or post-acute sequelae of SARS-CoV-2 (PASC)
  - **COVID-19 Negative Control Cohort**: No record of SARS-CoV-2 infection, ≥1 negative COVID-19 test; index date randomly sampled from the empirical distribution of infection dates in COVID-19 positive cohort
- **Outcome of interests**:
  - **Dyslipidemia**:
    - Abnormal TC: Total cholesterol (TC) ≥ 200 mg/dL
    - Abnormal TG:
      - Triglycerides (TG) ≥ 100 mg/dL (ages 0-9 years)
      - ≥ 130 mg/dL for (ages 10-19 years)
      - ≥ 150 mg/dL for (ages 20-21 years)
    - Abnormal LDL: Low-density lipoprotein (LDL) cholesterol ≥ 130 mg/dL
    - Abnormal HDL: High-density lipoprotein (HDL) cholesterol < 40 mg/dL
    - Abnormal non-HDL: Non-HDL cholesterol ≥ 145 mg/dL

Note: we removed Abnormal Apolipoprotein B: Apolipoprotein B ≥ 110 mg/dL due to an overall incidence below 0.1%.

- Abnormal BMI:
  - BMI z-score≥ 95th percentile (ages 2-18 years)
  - BMI ≥ 30 kg/m^2^ (ages 19-21 years)
- **Baseline Period (washout & covariate capture)**
  - Window: 7-729 days before index
  - Required ≥1 clinical encounter (inpatient, outpatient, or ED)
  - Excluded if any record of the outcome of interest during this period
  - **Additional Exclusions (Abnormal-BMI cohorts only)**:
    - Pre-existing abnormal conditions:
      - Cancer, cystic fibrosis, eating disorder, sickle cell disease, Crohn’s disease, ulcerative colitis, HIV, growth hormone deficiency, Cushing syndrome, panhypopituitarism, BMI less then 5th percentile for age and sex, pregnancy, bariatric surgery
    - Weight-modifying medications:
      - metformin, orlistat, liraglutide, exenatide, dulaglutide, semaglutide, setmelanotide, phentermine, topiramate
- **Follow-Up Period (outcome ascertainment, post-acute phase)**
  - Window: 28–179 days after index
  - Required ≥1 clinical encounter

To guarantee a comprehensive follow-up for all participants, we mandated that entry into the cohort—applicable to both the exposure and control groups—be timed no later than 179 days prior to the conclusion of the study period. This stipulation was critical to secure a full 179-day follow-up duration for each participant, thereby upholding the integrity of the follow-up data.

#### D. Study variables

**Table S1**. Variables used in the study evaluating the relative risk of post-acute dyslipidemia and abnormal BMI outcomes after SARS-CoV-2 infection in children and adolescents.

| Variable | Functional form | Values | Detail | Codes/references |
| --- | --- | --- | --- | --- |
| Treatment (i.e., Exposure) |  |  |  |  |
| Documented SARS-CoV-2 infection | Indicator | Yes/No | Based on the observation and visit occurrence domains. Defined as a polymerase-chain-reaction (PCR), serology, or antigen tests positive for COVID-19, or diagnoses of COVID-19, post-acute sequelae of SARS-CoV-2 (PASC) regardless of the presence of symptoms. | See <https://github.com/PEDSnet/PASC/tree/main/observation_derivation_recover_ml_phenotype/specs> for detailed codes. |
| Outcome |  |  |  |  |
| Abnormal Total cholesterol (TC) | Indicator | Yes/No | TC ≥ 200 mg/dL | See <https://github.com/lyqlei/PASC-Dyslipid_BMI/blob/74d6b87071643ef513482ccaf70f5ed444cc29fd/codeset_measurement_table_dyslipid.csv> for detailed SNOMED concept codes. |
| Abnormal Triglycerides (TG) | Indicator | Yes/No | TG:   - ≥ 100 mg/dL, 0-9 years; - ≥ 130 mg/dL, 10-19 years; - ≥ 150 mg/dL, 20-21 years |  |
| Abnormal low-density lipoprotein (LDL) cholesterol | Indicator | Yes/No | LDL cholesterol ≥ 130 mg/dL |  |
| Abnormal high-density lipoprotein (HDL) cholesterol | Indicator | Yes/No | HDL cholesterol < 40 mg/dL |  |
| Abnormal non-HDL cholesterol | Indicator | Yes/No | Non-HDL ≥ 145 mg/dL |  |
| Any abnormal lipid laboratory result | Indicator | Yes/No | Including incident occurrence of any dyslipidemia outcome studied. |  |
| Abnormal BMI | Indicator | Yes/No | - BMI z-score ≥ 95^th^ percentile, 2-18 years; - BMI ≥ 130 mg/dL, 19-21 years | Measurement concept ID: 3038553 for BMI, 2000000043 for BMI z-score |
| Confounding variables |  |  |  |  |
| Age (years) | Linear | NA | Based on records in the person domain. | Age is defined as the integer of (date – birth date)/365.25 |
| Sex | Indicator | Male/Female | Based on records in the person domain. | NA |
| Race/Ethnicity | 6 categories | NHW  NHB  Hispanic  AAPI  Multiple  Other/unknown | Based on records in the person domain. | NA |
| Obesity | 3 categories | Yes/No/Unknown | Based on records in the measurement domain.  If measured at age < 24*30.5 days, NHANES weight z score > 1.64  If measured at 24*30.5 < age <240*30.5, NHANES BMI z score > 1.64  If measured at age >= 240*30.5, BMI kg/m2 > 30 | NA |
| PMCA (Pediatric Medical Complexity Algorithm) | 3 categories | No chronic condition (PMCA = 0)  Non-complex chronic condition (PMCA = 1)  Complex chronic condition comorbidities (PMCA = 2) | Based on the condition occurrence and visit occurrence domains. | ^1^ |
| Diagnosis of each chronic condition cluster in 24 months to 7 days prior to the entry | Indicator | Yes/No | 205 chronic condition clusters were defined based on the condition occurrence and visit occurrence domains. | ^2^ |
| Number of visits to emergency department in 24 months to 7 days prior to the entry | 4 categories | 0/1/≥2 | Based on the condition occurrence and visit occurrence domains. | NA |
| Number of inpatient visits in 24 months to 7 days prior to the entry | 4 categories | 0/1/≥2 | Based on the condition occurrence and visit occurrence domains, including Inpatient Hospital Stay, Emergency Department Admit to Inpatient Hospital Stay, and Observation Stay | NA |
| Number of outpatient visits in 24 months to 7 days prior to the entry | 4 categories | 0/1/≥2 | Based on the condition occurrence and visit occurrence domains including Ambulatory/Outpatient Visit (With a Physician) and Interactive Telemedicine Service | NA |
| Number of unique medications in 24 months to 7 days prior to the entry | 4 categories | 0/1/≥2 | Based on the drug exposure domain. | NA |
| Number of negative COVID-19 tests in 24 months to 7 days prior to the entry | 4 categories | 0/1/≥2 | Based on the observation derivation recover domain | NA |
| Stratification variable |  |  |  |  |
| Obesity Status |  | Healthy (non-obese), Obesity class 1, Obesity class 2, Obesity class 3 | Healthy:   - BMI z-score ≥ –1.645 and < 1.036 (5^th^ to 85^th^ percentile), 0-19 years; - 18.5 ≤ BMI < 25, 20-21 years   Obesity class 1:   - BMI z-score > 1.645 (95^th^ percentile), 0-19 years; - 30 ≤ BMI < 35, 20-21 years   Obesity class 2:   - 120% of 95^th^ percentile for sex/age ≤ BMI < 140% of 95^th^ percentile for sex/age, 0-19 years - 35 ≤ BMI < 40, 20-21 years   Obesity class 3:   - BMI ≥ 140% of 95^th^ percentile for sex/age -OR- BMI ≥ 40, 0-19 years - BMI ≥ 40, 20-21 years | BMI z-score classification was based on age- and sex-specific CDC 2000 growth charts for participants aged 2 to 19 years |
| Other variables for eligibility criteria |  |  |  |  |
| Prior encounter in 24 months to 7 days prior to the entry | Indicator | Yes/No | Based on the condition occurrence and visit occurrence domains. | NA |
| Follow-up encounter in 28 to 179 days after the entry | Indicator | Yes/No | Based on the condition occurrence and visit occurrence domains. | NA |

Note: All the domains in the table above are based on the PEDSnet common data model (CDM). More details are available through this link: <https://data-models-service.research.chop.edu>.

### Section S2 Supplemental Results: patient characteristic balance between COVID-19 positive and negative groups

#### A. Propensity-score (PS) models and stratification

We fitted a large-scale PS model for the study cohort with baseline patient characteristics including

- Demographics (age at index date; sex; race/ethnicity)
- Obesity (Yes/No/Unknown)
- Chronic condition indicator as defined by the Pediatric Medical Complexity Algorithm (PMCA)
  - No chronic condition (PMCA = 0)
  - Non-complex chronic condition (PMCA = 1)
  - Complex chronic condition comorbidities (PMCA = 2)
- The existence of a list of 205 chronic conditions 24 months ~ 7 days prior to the index
- Healthcare utilization 24 months ~ 7 days prior to index date categorized to 0,1,2, ≥3
  - Number of inpatient visits
  - Number of outpatient visits
  - Number of emergency department (ED) visits
  - Number of unique medications
  - Number of negative COVID-19 tests
- Cohort entry date (index date) categorized to 1 month
- Healthcare system index

The patients were then stratified into 6 equally spaced PS strata.

#### B. Indication Bias for Dyslipidemia Analysis

Lipid testing is not routinely conducted in pediatric practice, children who underwent these evaluations may differ systematically from those who did not. For example, patients who are more likely to complete the lipid lab test may also be more likely to have underlying health conditions that predispose them to abnormal lipid levels. Completing the lipid lab test may be associated with both exposure (COVID-19 infected or not) and outcome (abnormal lipid lab results). To address this, we further included a baseline indicator for participating lipid test in our covariates set^3,4^. All confounders were balanced after propensity score stratification, with an SMD of less than 0.1, **Figure S2**.

#### C. Empirical equipoise assessment

To assess the similarity across study groups, we present the preference score. This metric refines the propensity score by integrating treatment prevalence, facilitating an easily understood comparison. The preference score (F) is mathematically derived from the propensity score (S) and the treatment prevalence (P) using the following formula^5,6^:

$$\ln\left( \frac{F}{1-F} \right)=\ln\left( \frac{S}{1-S} \right)-\ln\left( \frac{P}{1-P} \right).$$

**Figure S1** below illustrates the preference score distributions for both treatment groups within each study cohort, which indicates the high comparability of these studies.

**Figure S1(a-g)**. Distribution of preference scores for COVID-19 positive and negative groups in each study populations. A greater convergence of these distributions indicates a higher similarity in the predicted likelihood of being infected between the COVID-19 positive (target cohort, in red) and negative (comparator cohort, in blue) participants.

**Figure S1(a) Distribution of preference scores for studying abnormal HDL outcome**

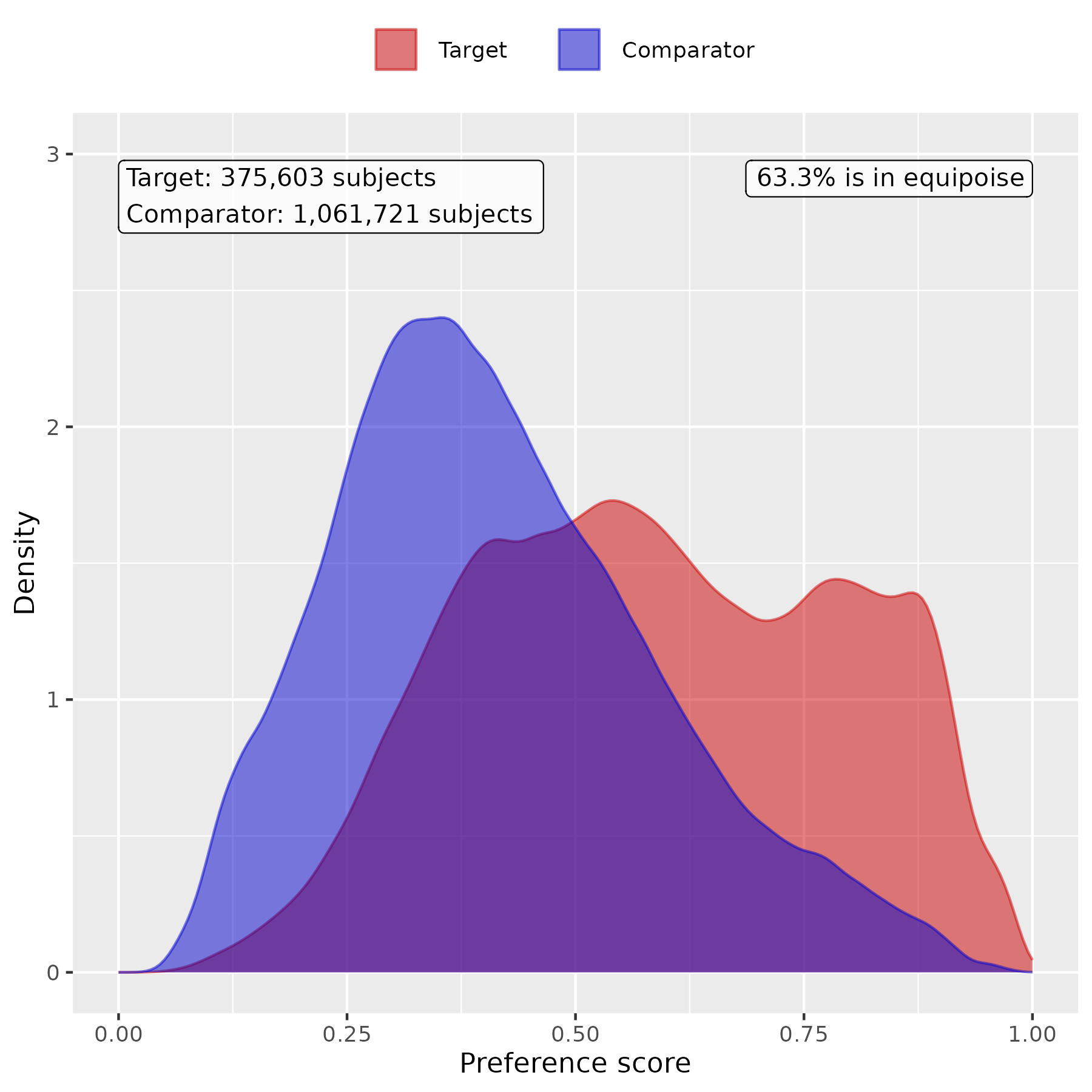

**Figure S1(b) Distribution of preference scores for studying abnormal LDL outcome**

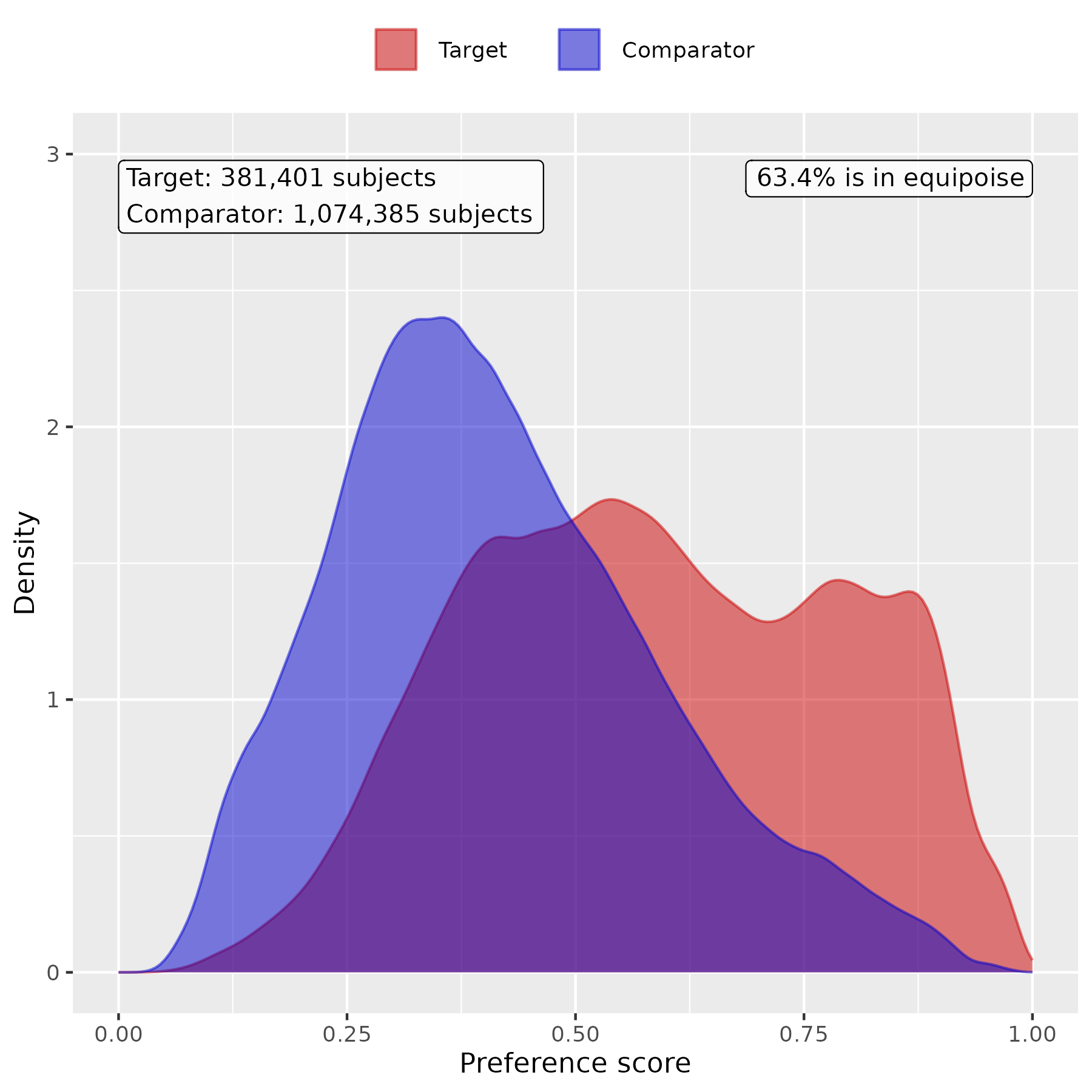

**Figure S1(c) Distribution of preference scores for studying abnormal non-HDL outcome**

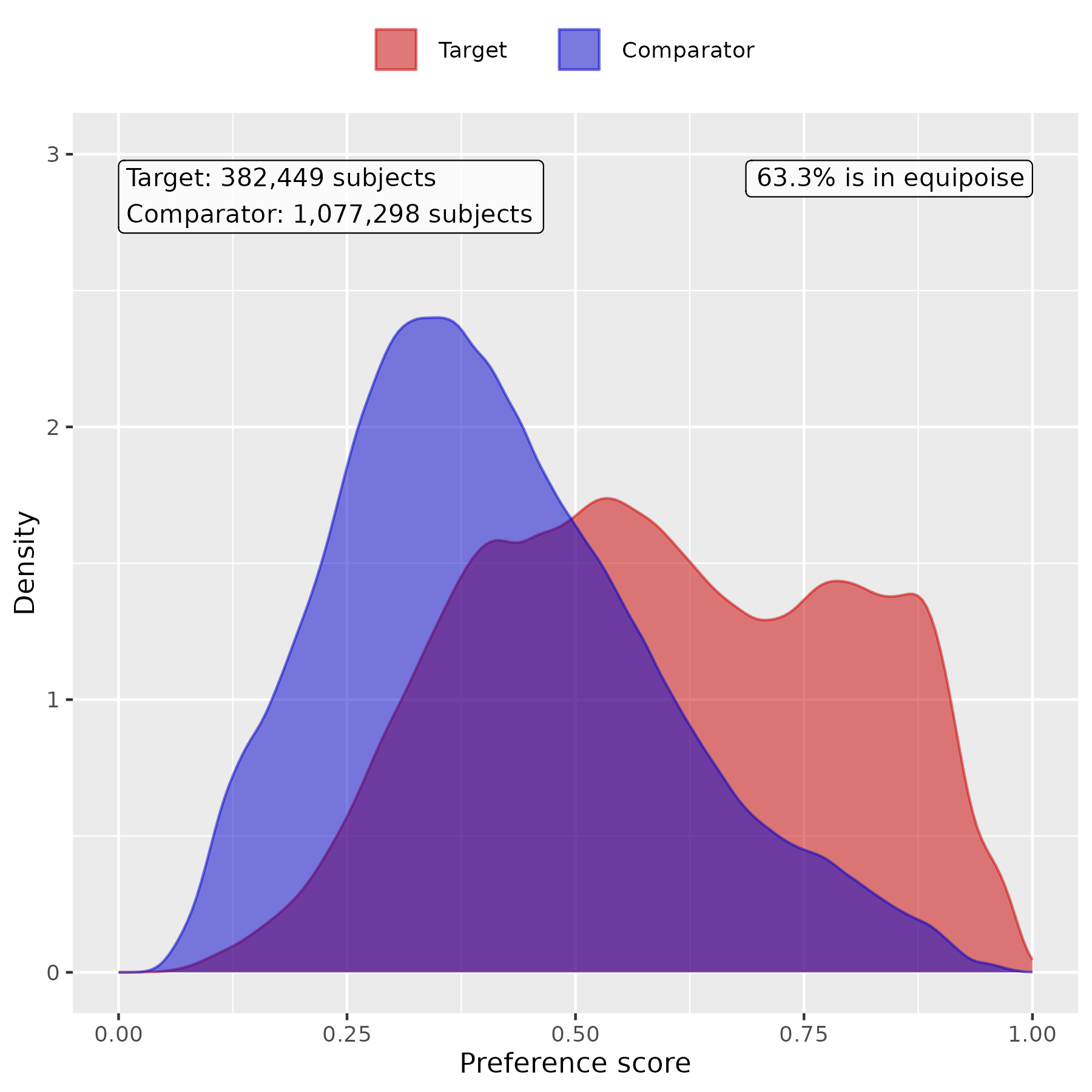

**Figure S1(d) Distribution of preference scores for studying abnormal TC outcome**

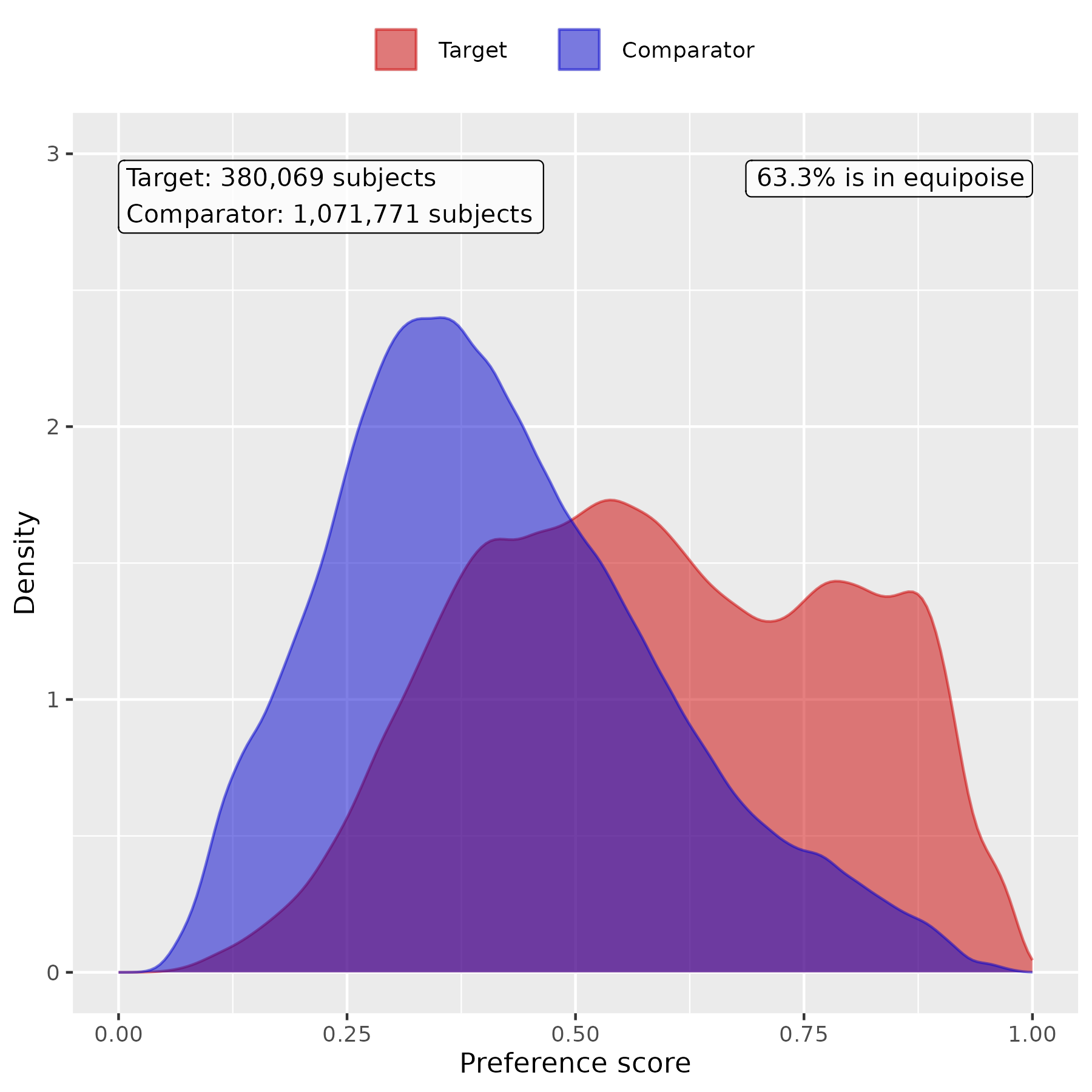

**Figure S1(e) Distribution of preference scores for studying abnormal TG outcome**

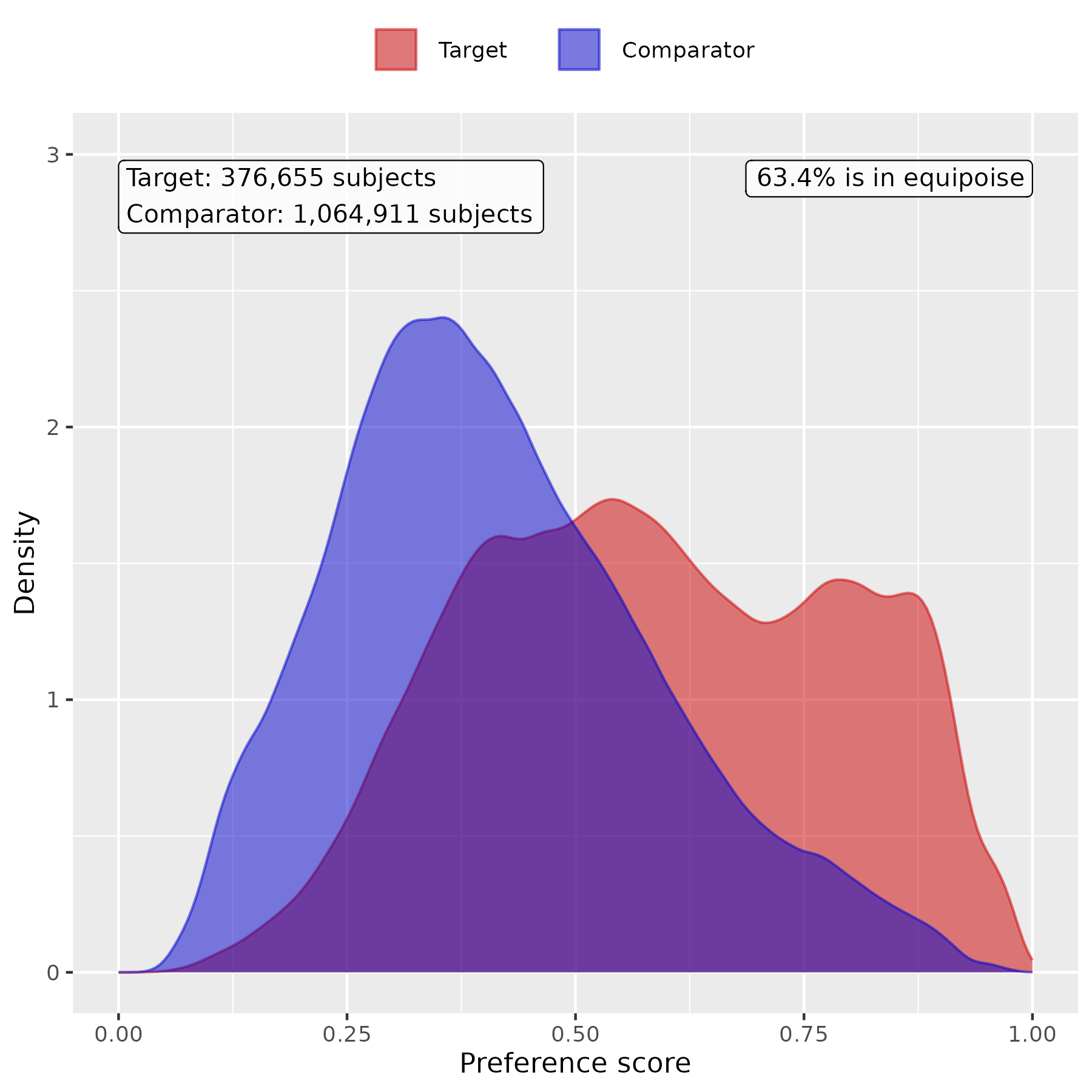

**Figure S1(f) Distribution of preference scores for studying any abnormal lab results outcome**

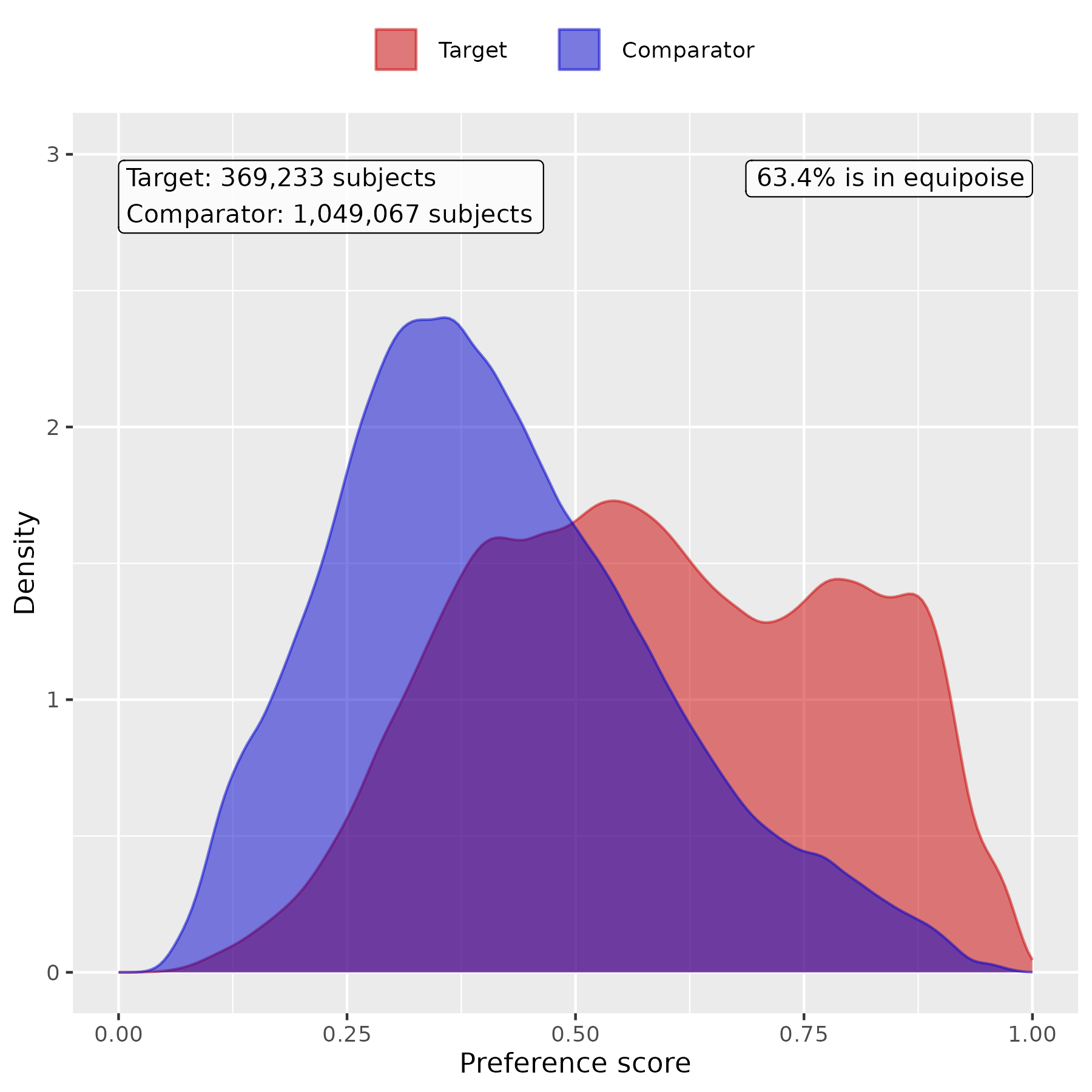

**Figure S1(g) Distribution of preference scores for studying abnormal BMI outcome**

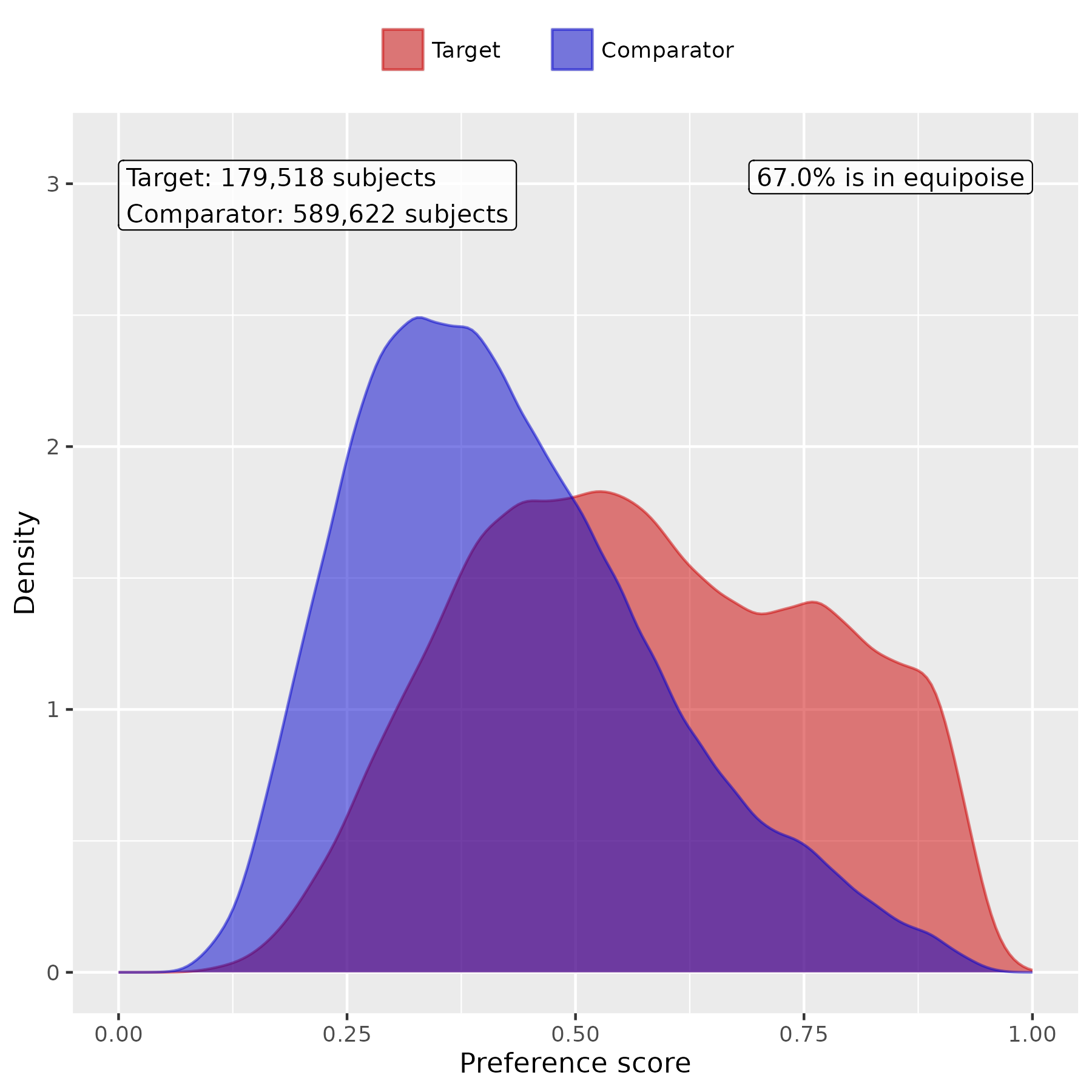

#### C. Patient characteristic balance in the primary analysis

We evaluate the balance of patient characteristics using the standardized mean difference (SMD). **Figure S2** presents the SMD of the study cohorts before and after PS score stratifications.

**Figure S2(a-g)**. Patient characteristic balance before and after large-scale PS stratification in each study population. The upper panel displays the top 20 covariates with the largest SMDs before stratification, while the lower panel displays the top 20 covariates with the largest SMDs after stratification.

**Figure S2(a) Patient characteristic balance before and after large-scale PS stratification for studying abnormal HDL outcome**

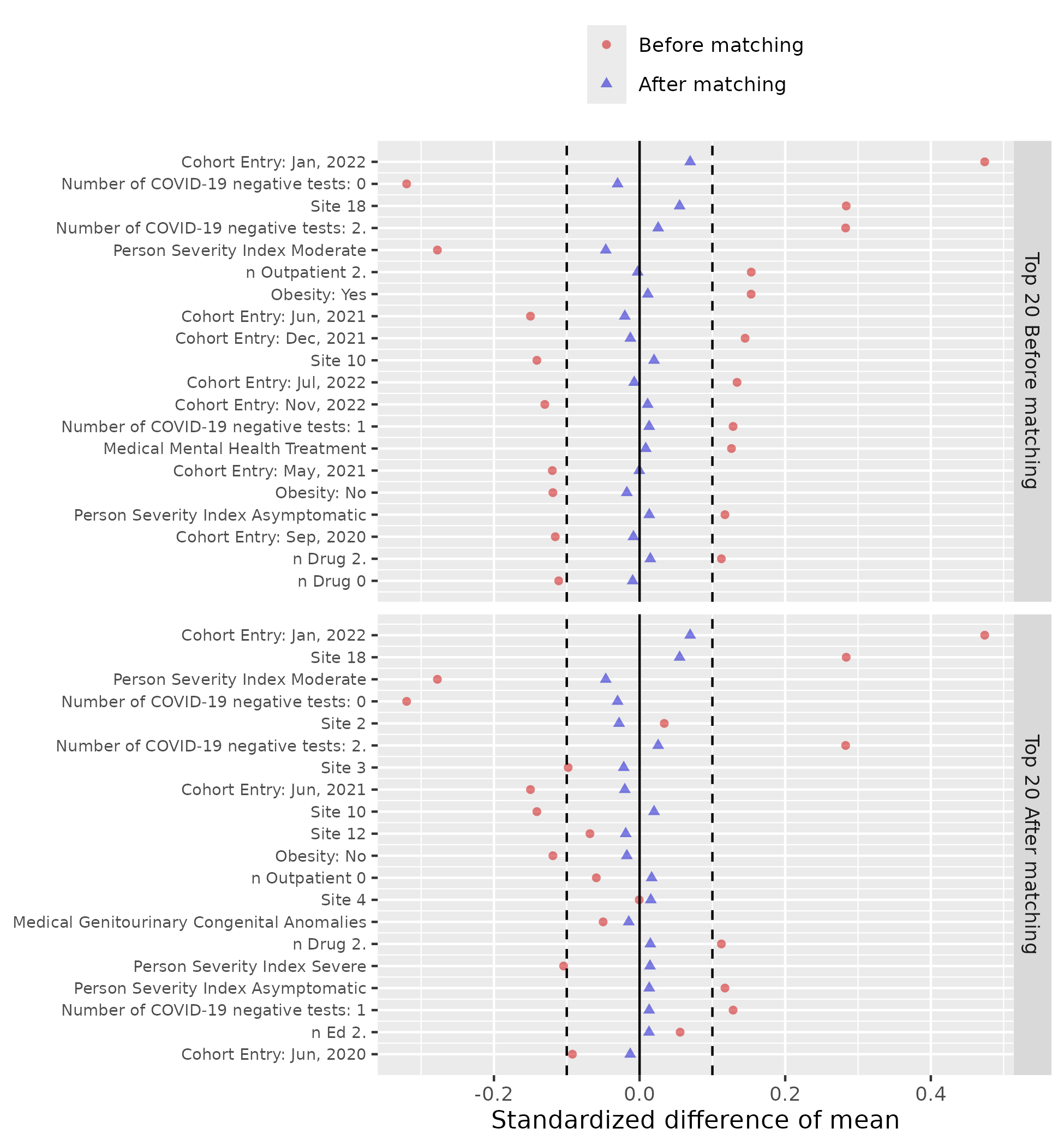

**Figure S2(b) Patient characteristic balance before and after large-scale PS stratification for studying abnormal LDL outcome**

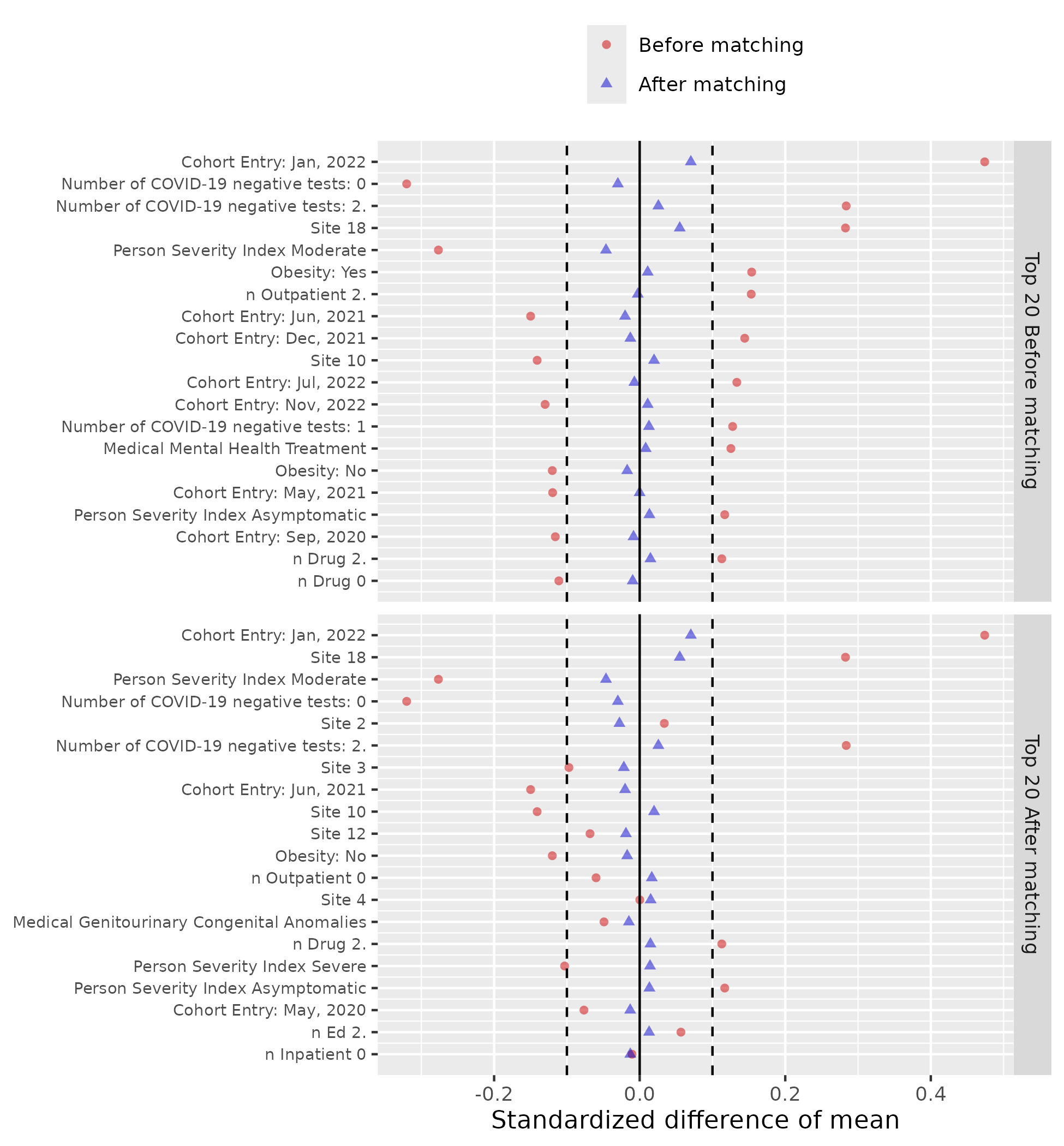

**Figure S2(c) Patient characteristic balance before and after large-scale PS stratification for studying abnormal non-HDL outcome**

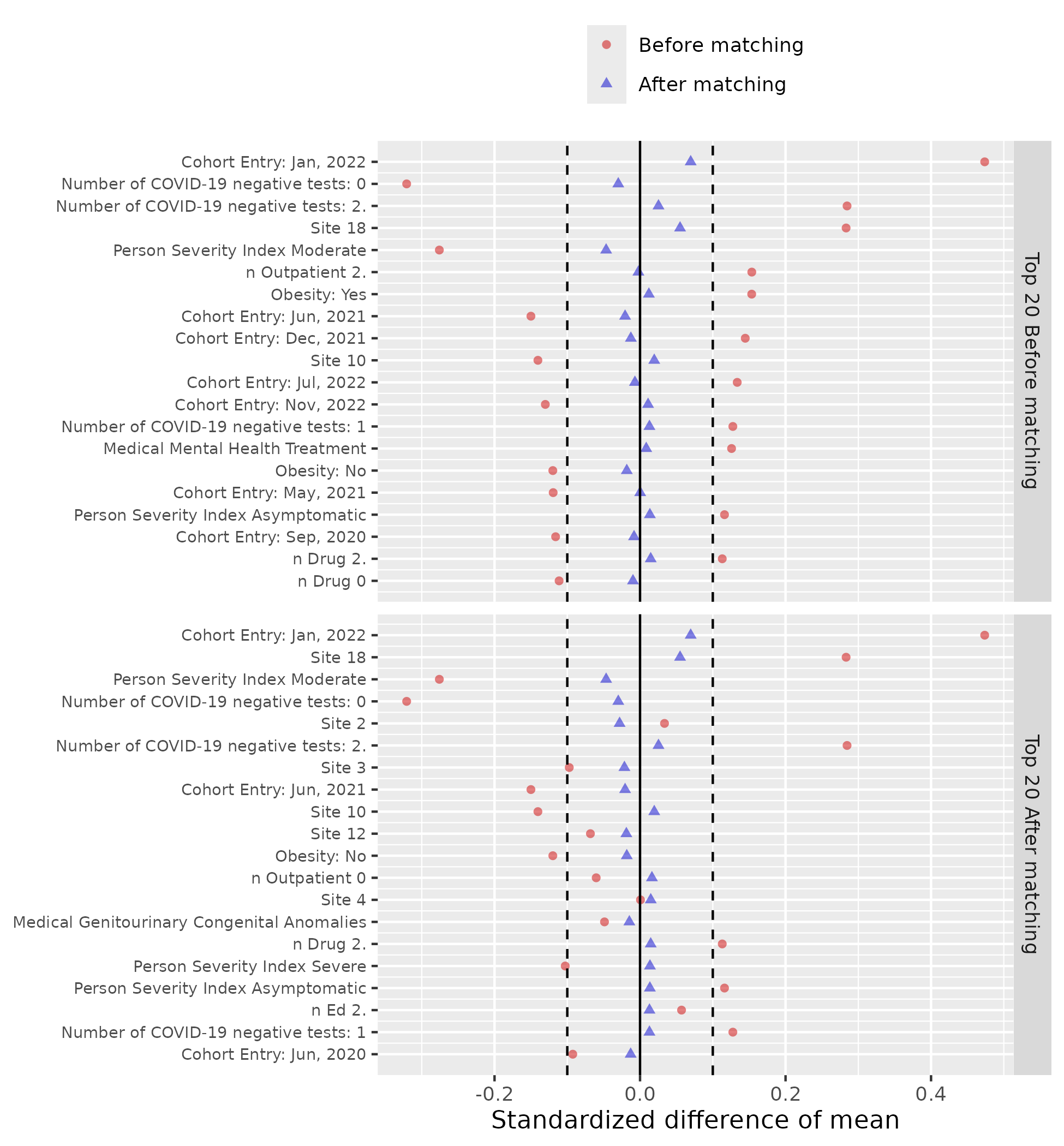

**Figure S2(d) Patient characteristic balance before and after large-scale PS stratification for studying abnormal TC outcome**

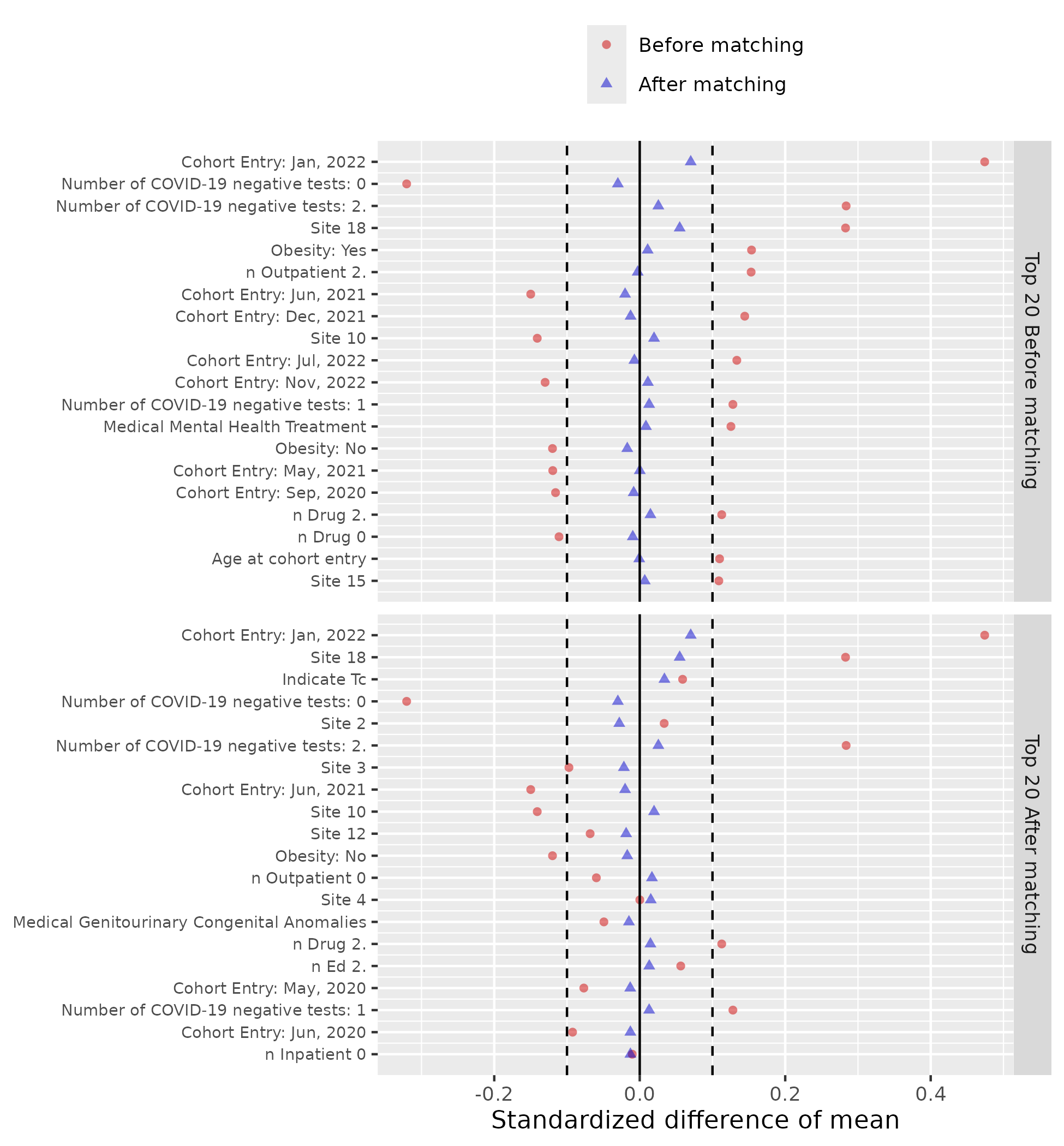

**Figure S2(e) Patient characteristic balance before and after large-scale PS stratification TG outcome**

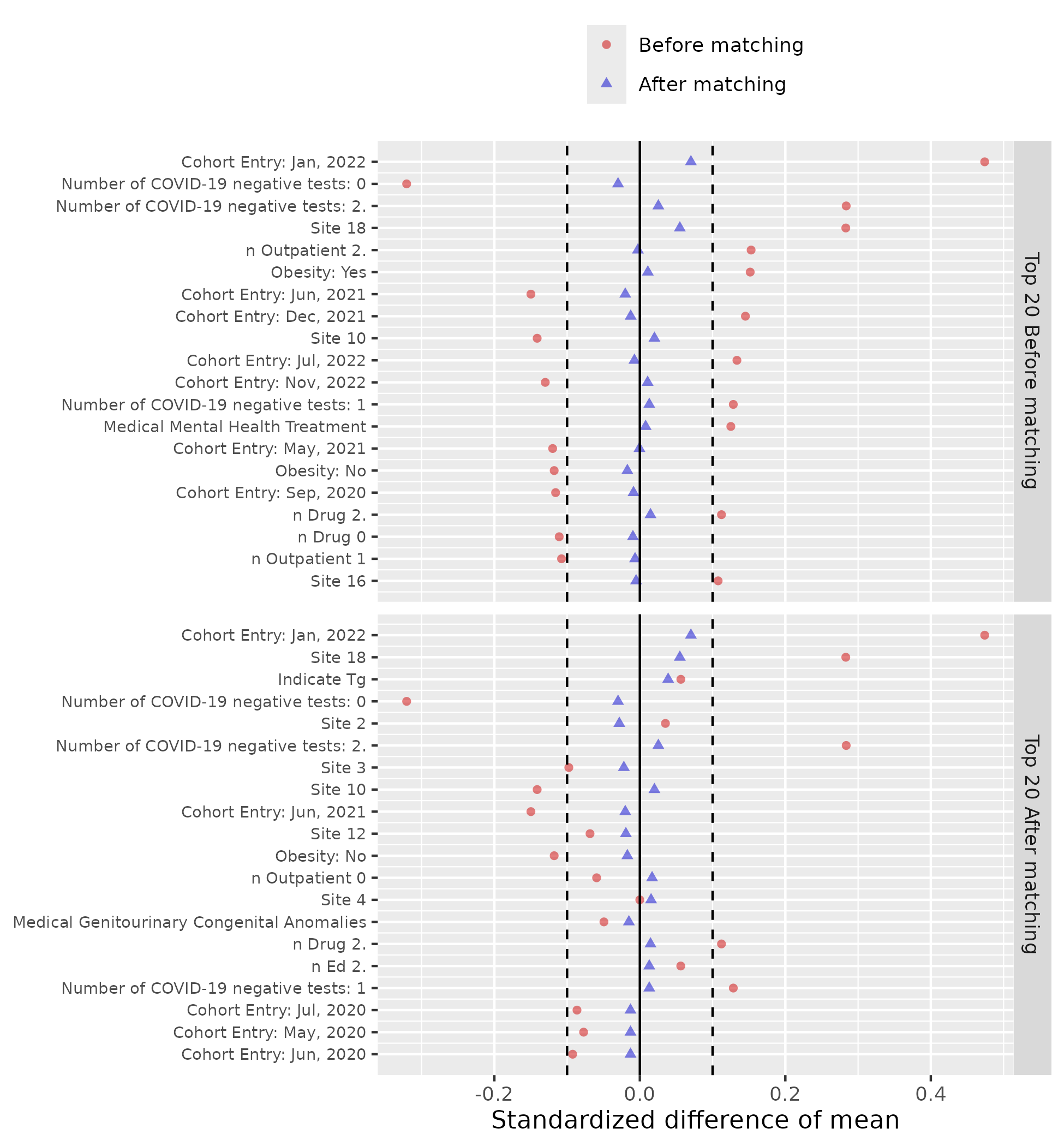

**Figure S2(f) Patient characteristic balance before and after large-scale PS stratification for studying any abnormal lab results outcome**

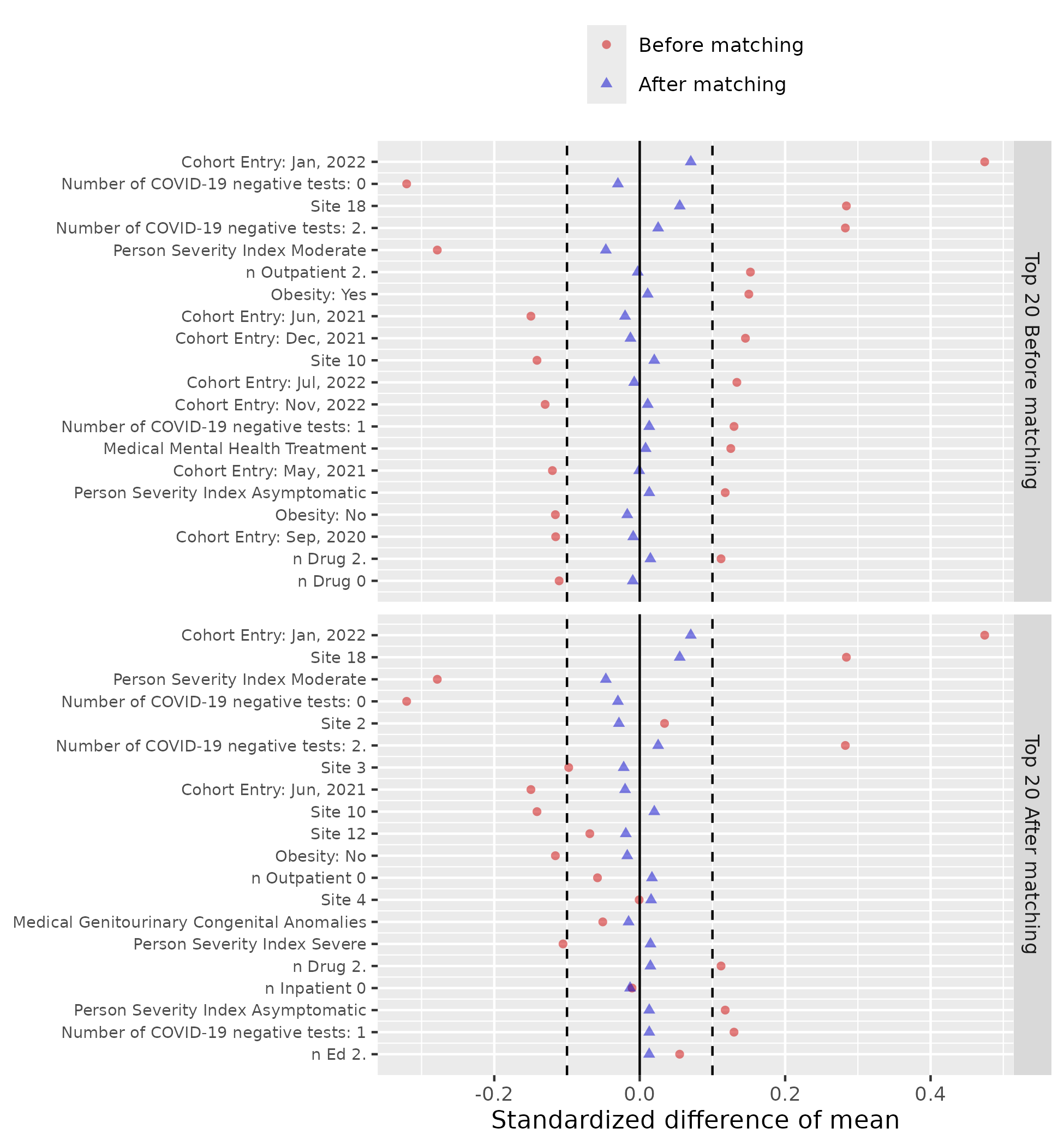

**Figure S2(g) Patient characteristic balance before and after large-scale PS stratification for studying abnormal BMI outcome**

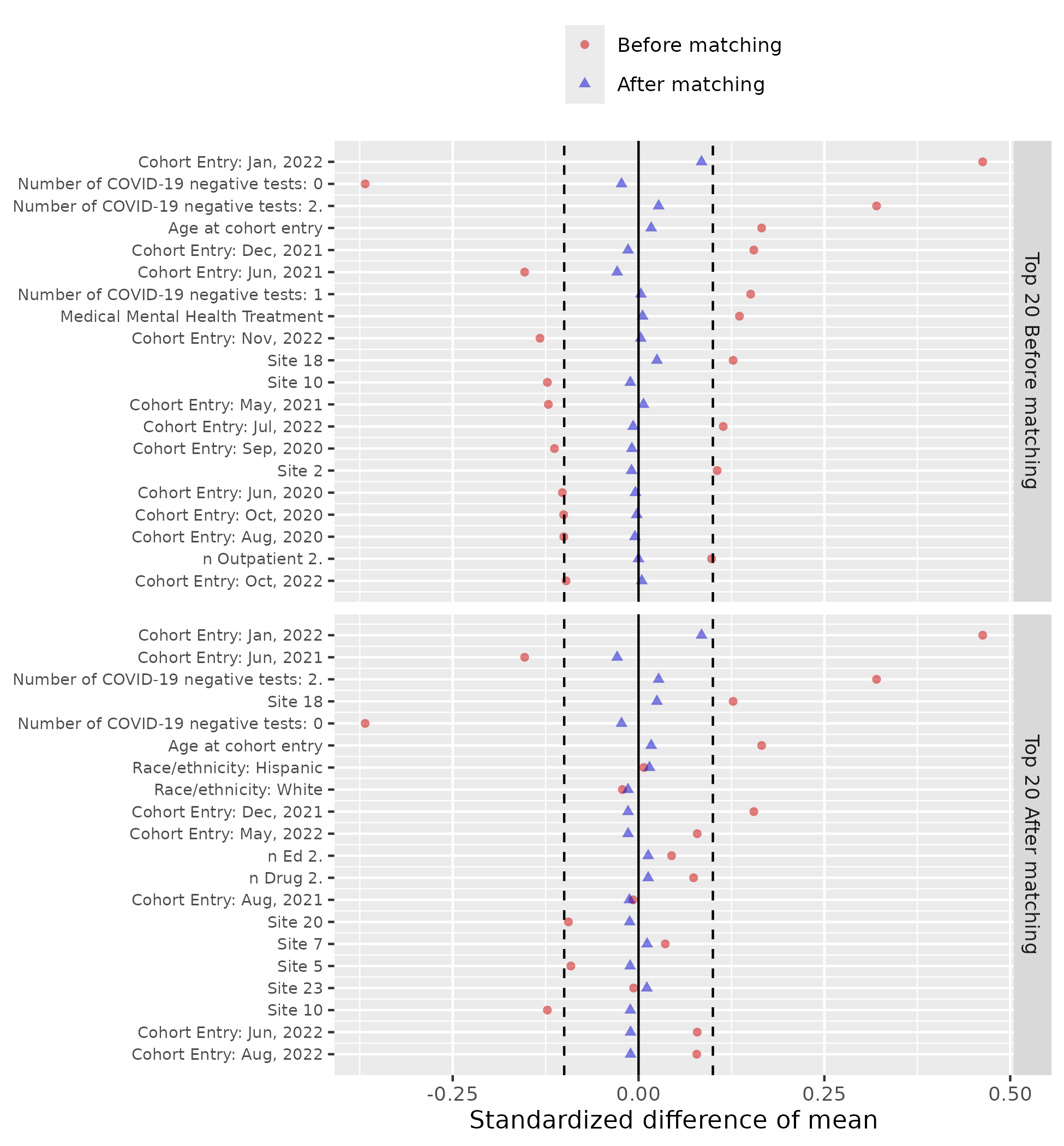

### Section S3 Supplemental Results: Negative Control Experiments

To evaluate the robustness of our findings, we conducted a series of negative control outcome (NCO) experiments using a predefined set of 36 outcomes. Negative control outcomes were defined as clinical conditions that are not plausibly causally related to the exposure of interest (COVID-19 in our analyses). The outcome list was developed in collaboration with pediatric clinicians based on domain knowledge and clinical judgment. A detailed enumeration of these outcomes is provided in Supplementary **Table S2**.

**Table S2**. List of negative control outcomes.

Health Conditions

Acne

Astigmatism

Autism/Autistic disorder

Closed fracture of distal end of radius

Closed injury of head

Concussion

Contact dermatitis

Diaper rash

Displacements - bone

Epilepsy

Falls

Foreign body in ear

Impetigo

Inguinal hernia

Injury of finger

Injury of free lower limb

Injury of head

Injury of left leg

Injury of right foot

Injury of right hand

Injury of right leg

Injury of upper extremity

Insect bite

Myopia

Plagiocephaly

Scoliosis

Seizure

Snoring/Obstructive sleep apnea

Speech delay

Speech dysfunction

Sprain of ankle

Tinea capitis

Tinea corporis

Tongue tie

Umbilical hernia

Wax in ear/impacted cerumen

We employed a set of 36 NCOs to assess the presence of residual bias and support calibration of our estimated risk ratios (RRs). These outcomes were selected based on clinical judgment by two board-certified pediatricians (DT, CF) and were defined as conditions with no known or plausible causal association with SARS-CoV-2 infection. Under the null hypothesis, the exposure is not expected to influence the incidence of these outcomes.

To implement negative control calibration, we first estimated the empirical null distribution derived from the effect estimates of the negative control outcomes. This distribution was then used to adjust the RRs from the primary analysis to account for potential systematic error. The methodological framework for deriving and applying the empirical null follows the principles outlined by Schuemie et al.^7,8^.

To improve the stability of these estimates and avoid excessive uncertainty, negative control outcomes with a cohort-wide incidence below 0.1% were excluded from the calibration analysis. The estimated RRs and standard errors for the included NCOs are shown in **Figure S3**. We quantified the magnitude of systematic error using the expected absolute systematic error (EASE), calculated as the absolute difference between the log-transformed observed RR and the (assumed null) true RR. A minimal degree of systematic bias was detected, and the calibrated estimates are reported in **Figure S4**.

**Figure S3(a-g)**. Systematic error control when estimating RR in each study population. The plot displays RRs and their corresponding standard errors for each negative control outcome.

**Figure S3(a) Systematic error control when estimating RR for studying abnormal HDL outcome**

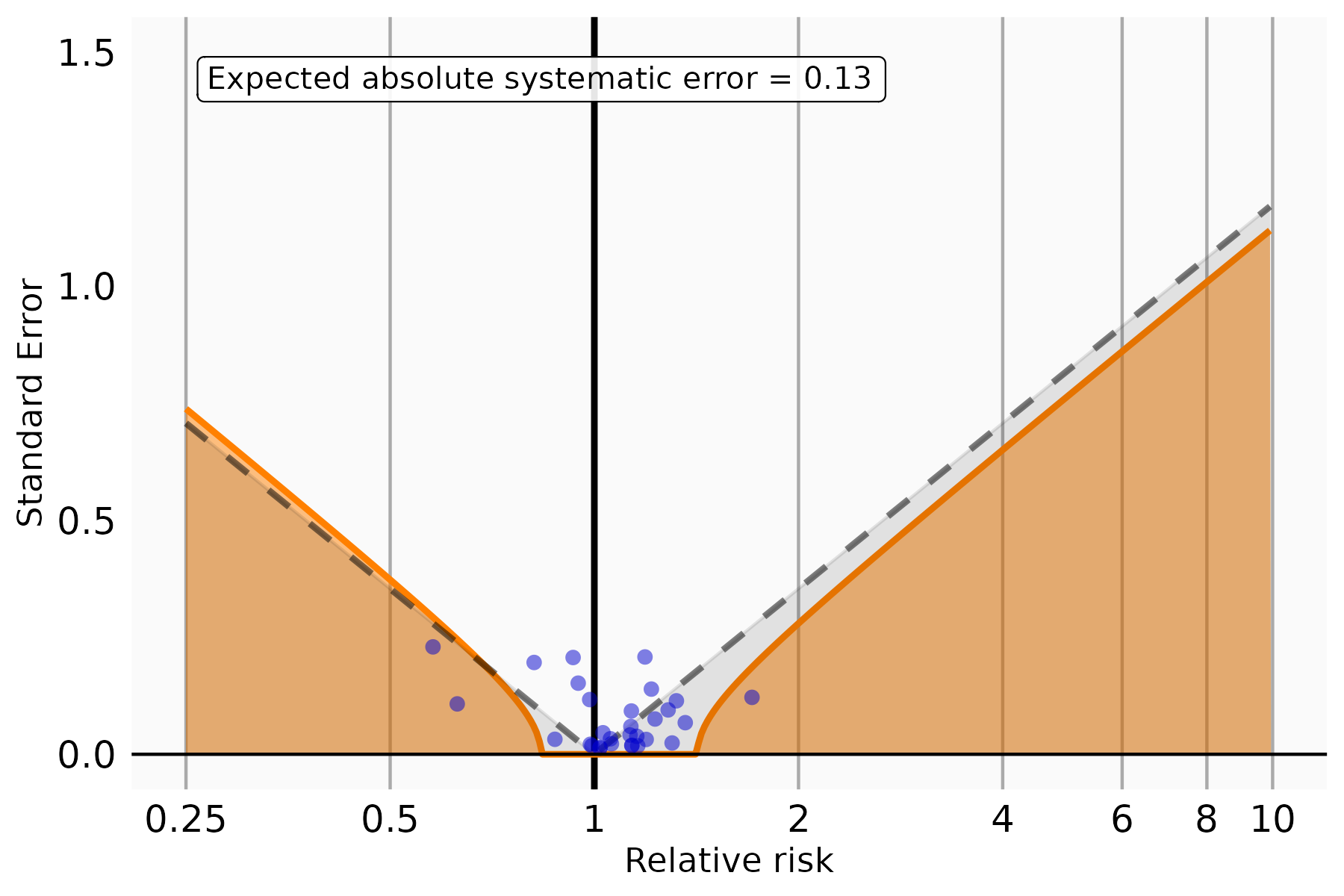

**Figure S3(b) Systematic error control when estimating RR for studying abnormal LDL outcome**

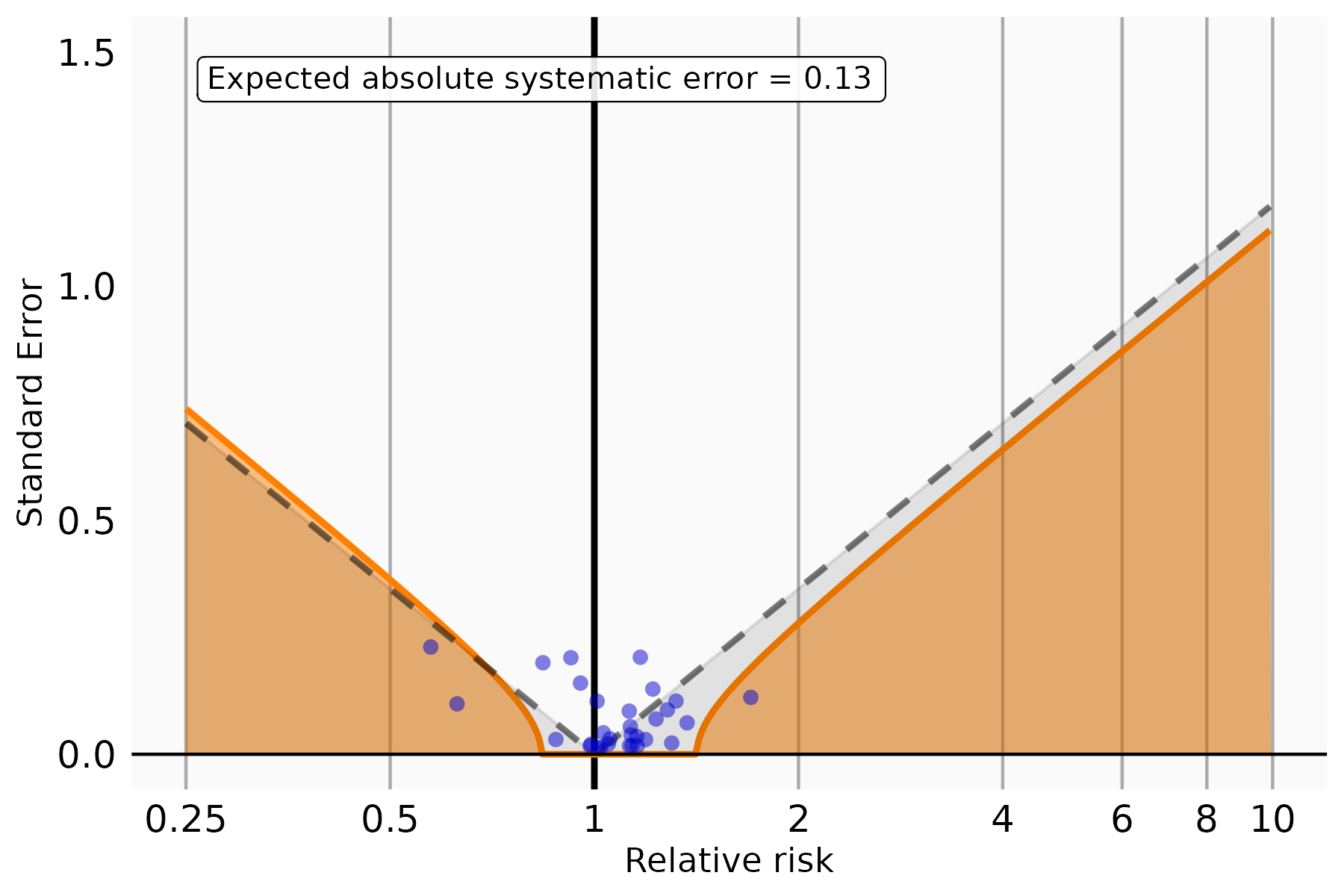

**Figure S3(c) Systematic error control when estimating RR for studying abnormal non-HDL outcome**

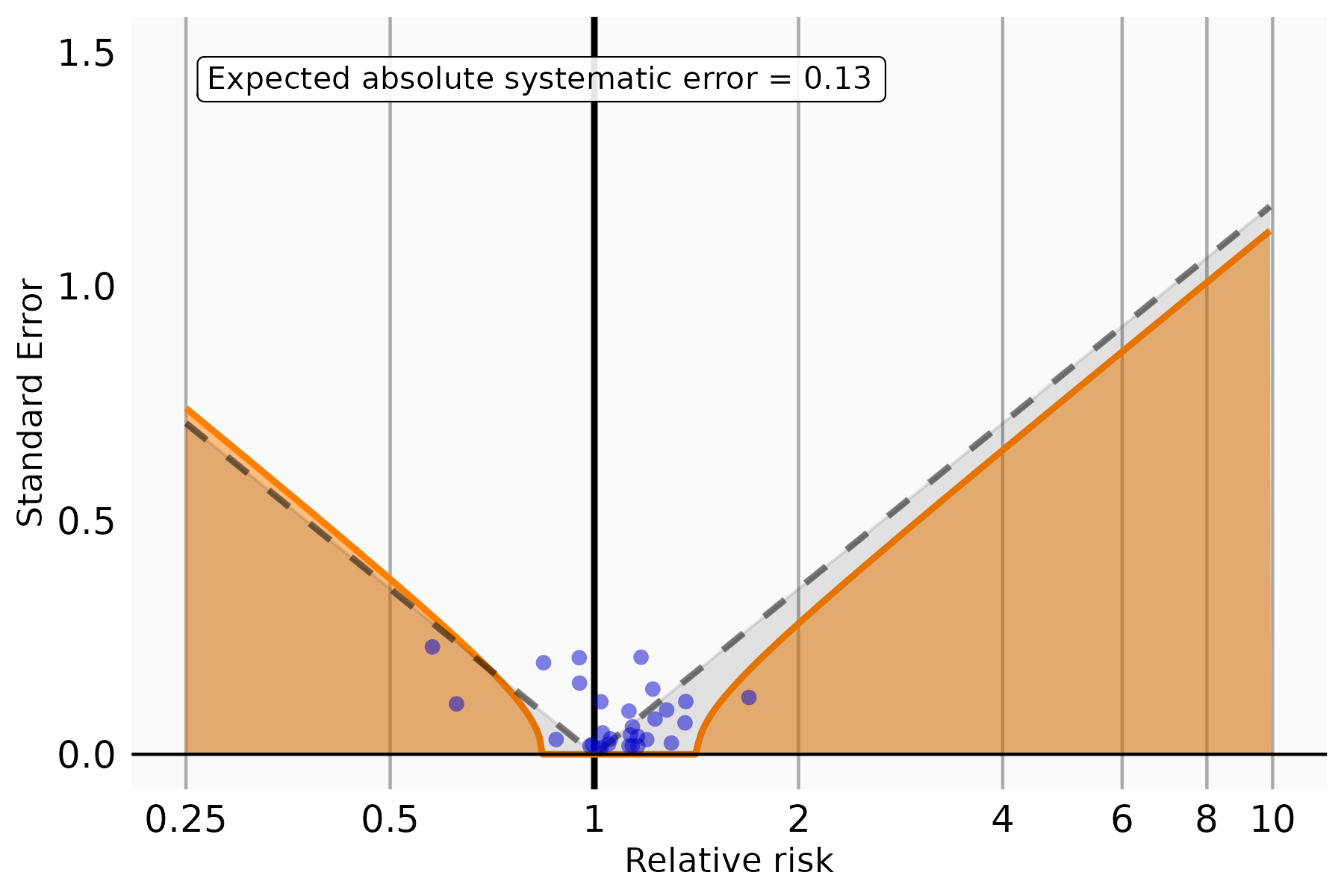

**Figure S3(d) Systematic error control when estimating RR for studying abnormal TC outcome**

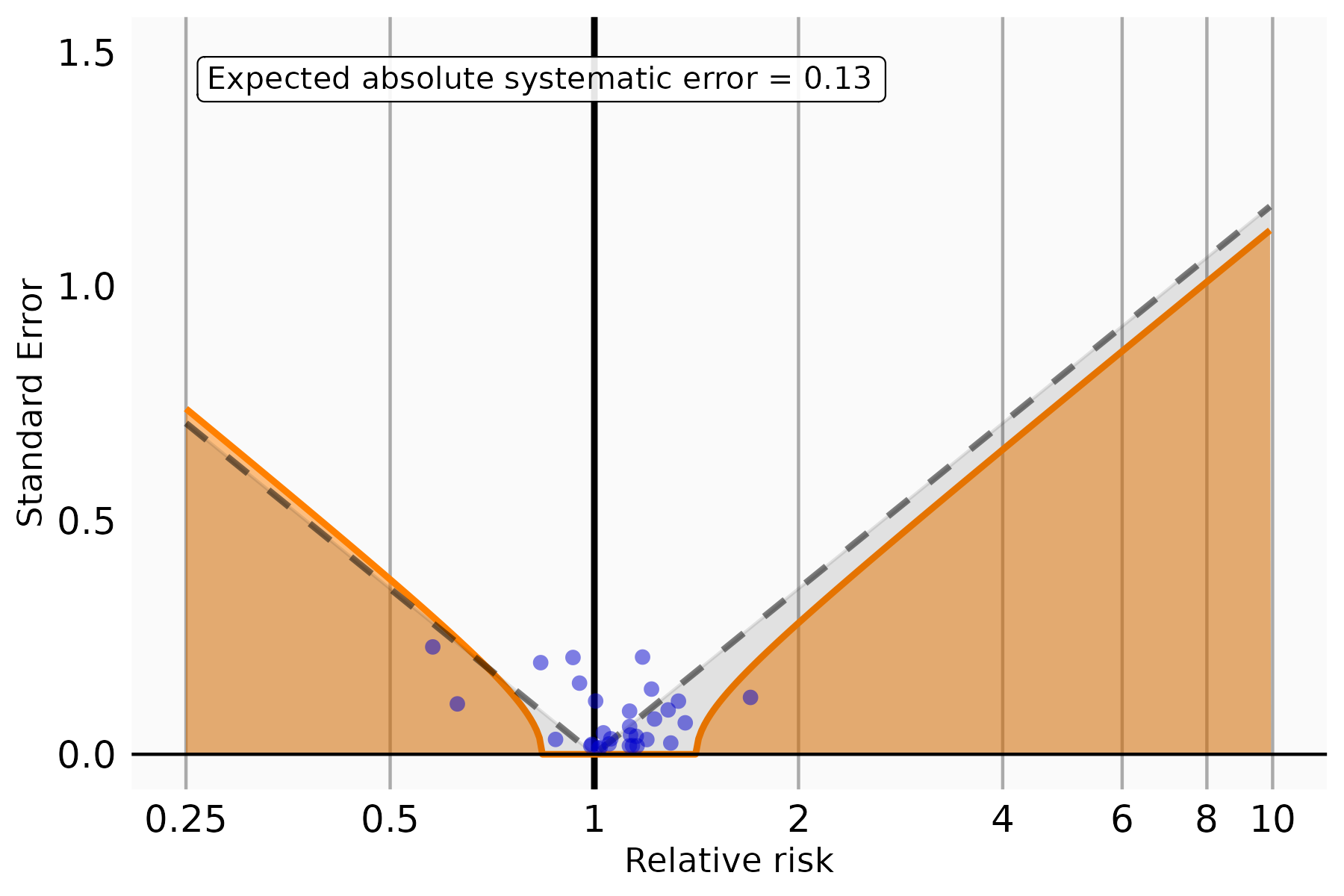

**Figure S3(e) Systematic error control when estimating RR for studying abnormal TG outcome**

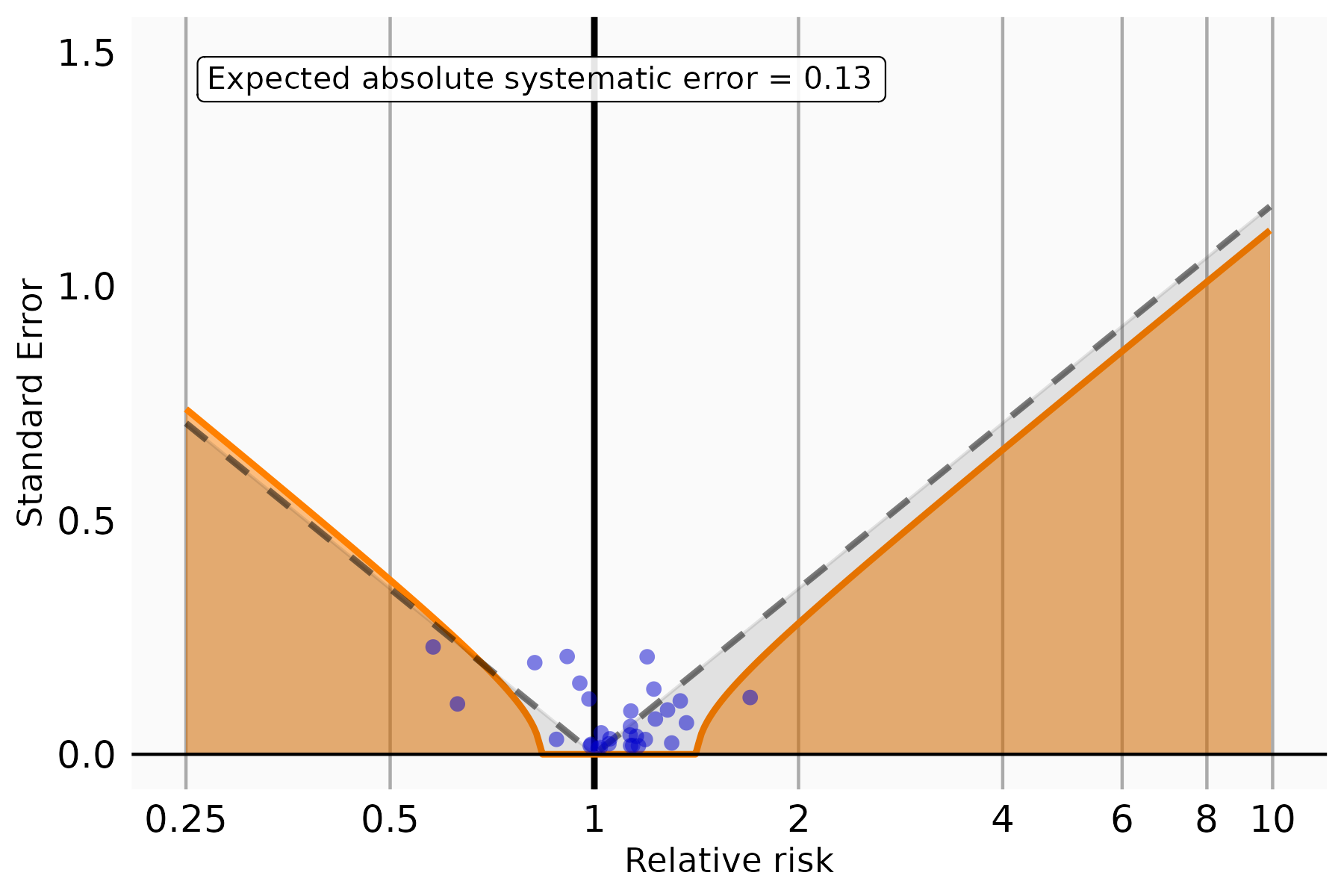

**Figure S3(f) Systematic error control when estimating RR for studying any abnormal lab results outcome**

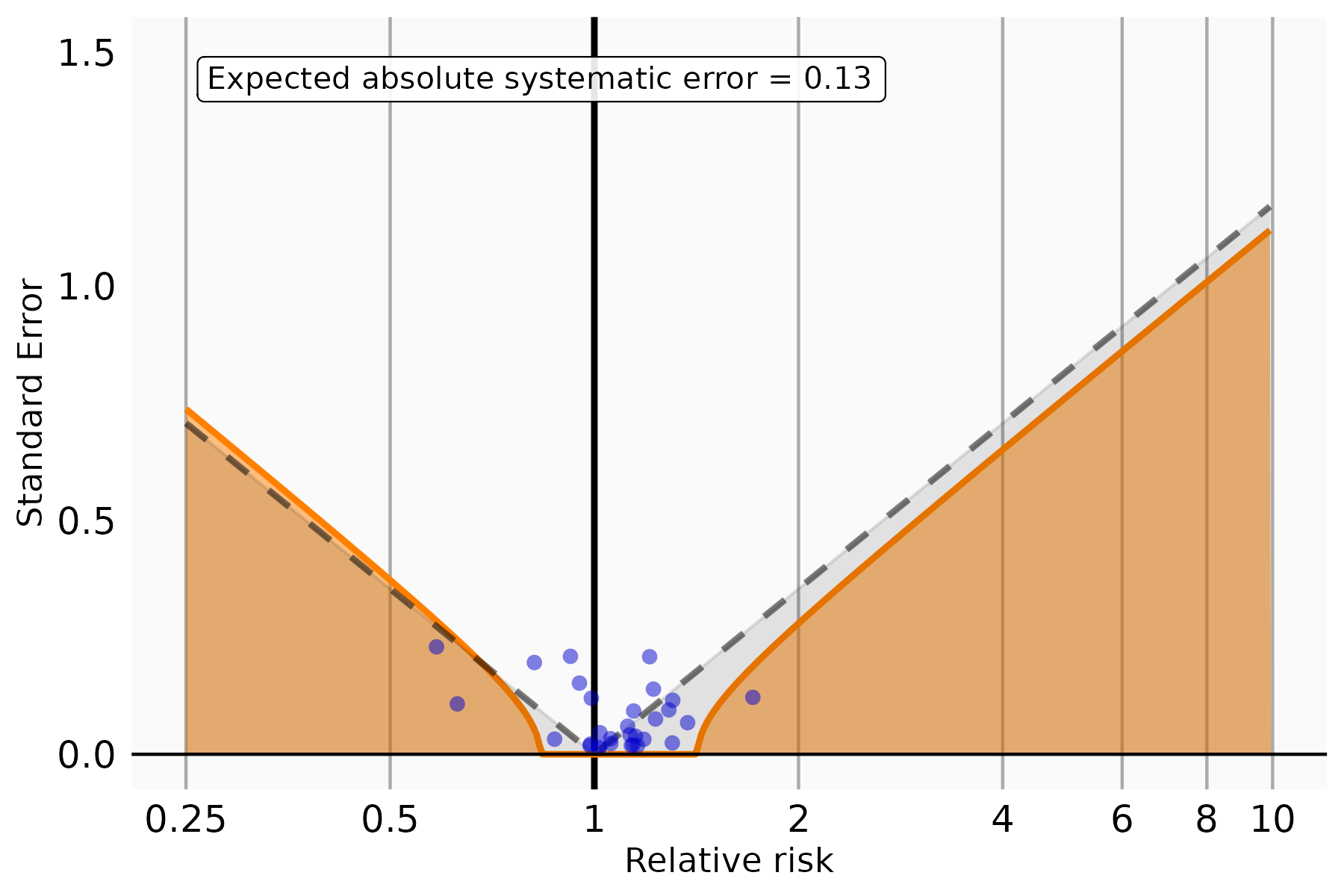

**Figure S3(g) Systematic error control when estimating RR for studying BMI outcome**

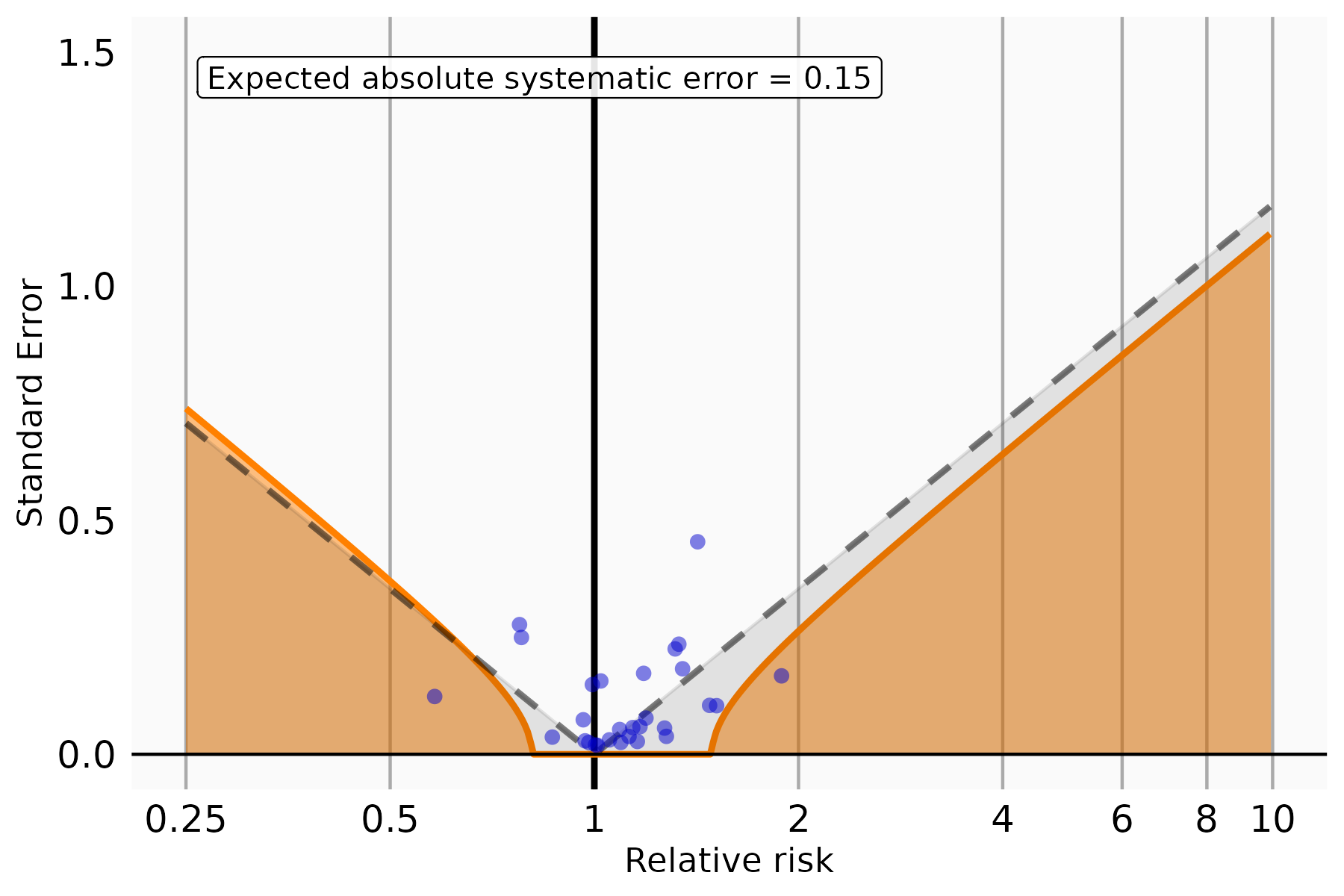

**Figure S4**. Risks of incident post-acute COVID-19 dyslipidemia and abnormal BMI outcomes with the contemporary control cohort, after negative control outcomes calibration.

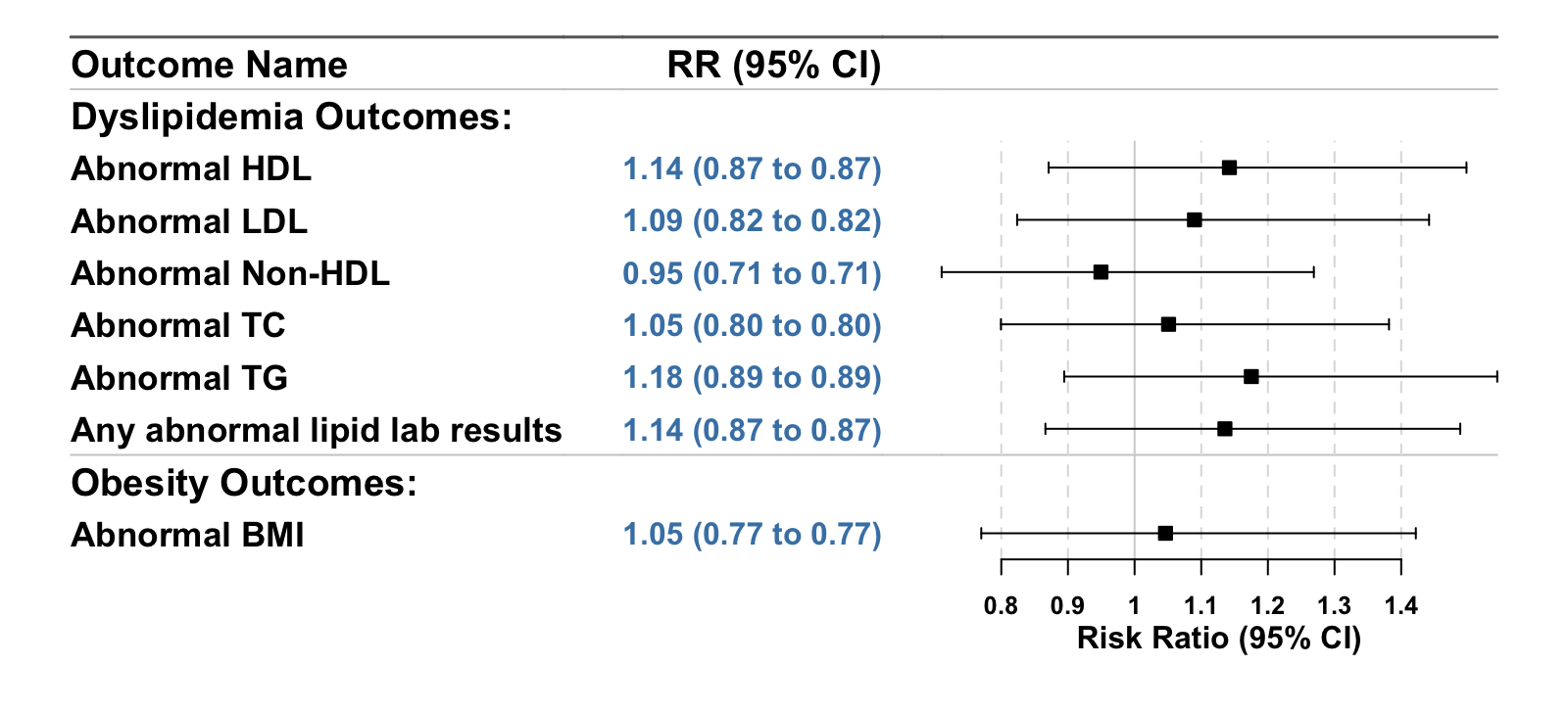

### Section S4 Supplemental Results: stratified analysis by obesity status subgroups

We conducted subgroup analyses for dyslipidemia outcomes stratified by baseline obesity status using age- and sex-specific BMI percentiles for individuals aged 0–19 years and standard BMI thresholds for those aged 20–21 years. Obesity categories were defined as follows:

- **Healthy weight**:
  - For participants aged 0–19 years: BMI z-score between –1.645 and < 1.036 (corresponding to the 5th to <85th percentiles)
  - For participants aged 20–21 years: BMI between 18.5 and <25 kg/m²
- **Obesity class 1**:
  - For participants aged 0–19 years: BMI z-score > 1.645 (≥95th percentile)
  - For participants aged 20–21 years: BMI between 30 and <35 kg/m²
- **Obesity class 2**:
  - For participants aged 0–19 years: BMI between 120% and <140% of the 95th percentile for age and sex
  - For participants aged 20–21 years: BMI between 35 and <40 kg/m²
- **Obesity class 3**:
  - For participants aged 0–19 years: BMI ≥140% of the 95th percentile for age and sex or absolute BMI ≥40 kg/m²
  - For participants aged 20–21 years: BMI ≥40 kg/m²

**Figure S5(a-c)**. Risks of incident post-acute COVID-19 dyslipidemia outcomes with the contemporary control cohort, by baseline obesity status subgroups.

**Figure S5(a) Overweight**

35,958 COVID-19 group v.s. 89,650 Negative control group

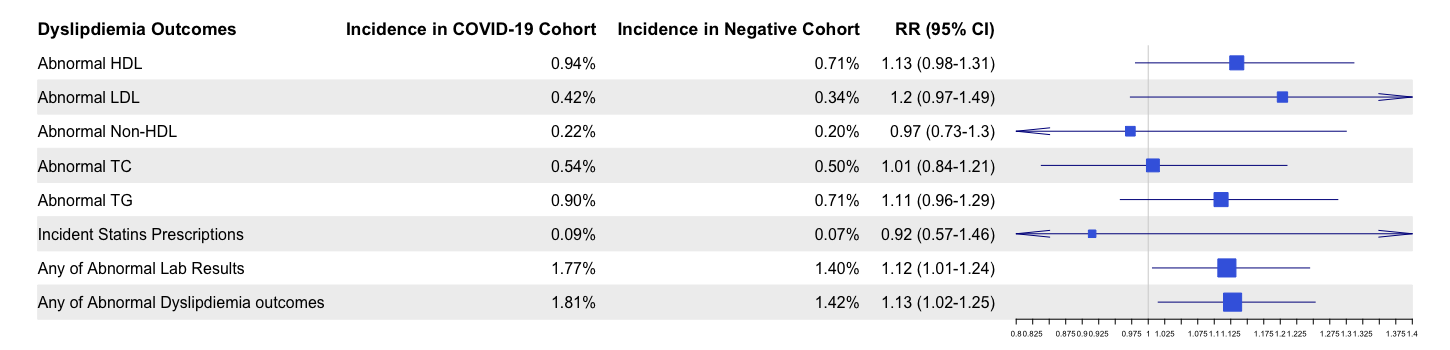

**Figure S5(b) Obesity (Class 1-3)**

384,289 COVID-19 group v.s. 1,080,413 Negative control group

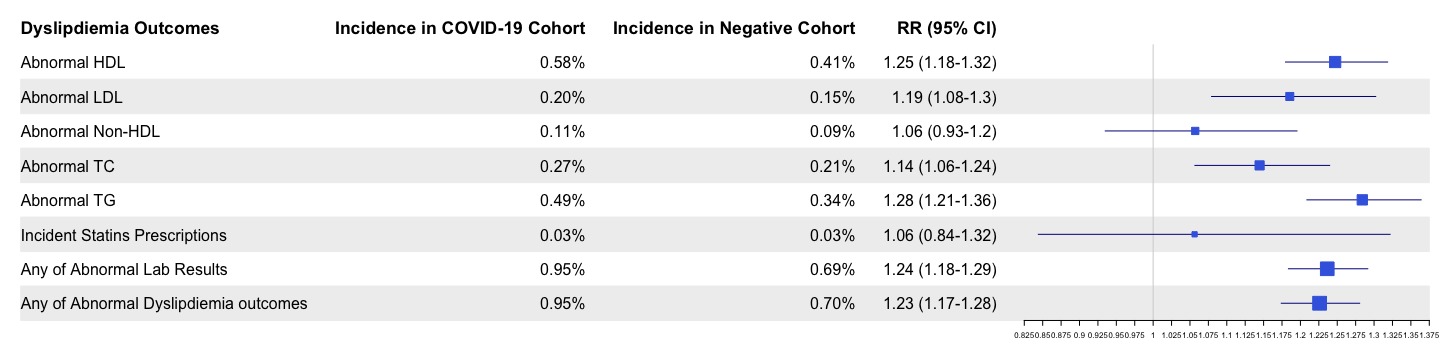

**Figure S5(c) Class 3 Obesity**

4,417 COVID-19 group v.s. 8,833 Negative control group

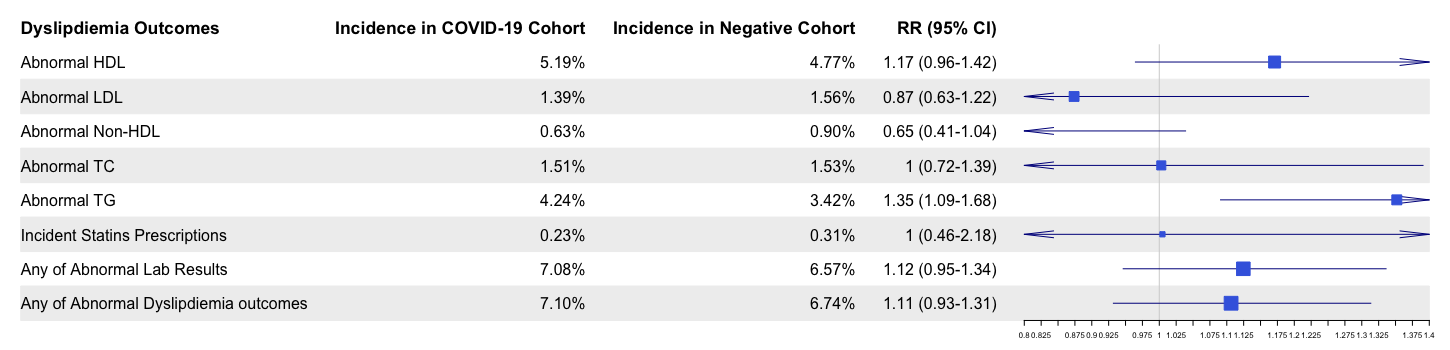
